# A foundation model for clinician-centered drug repurposing

**DOI:** 10.1101/2023.03.19.23287458

**Authors:** Kexin Huang, Payal Chandak, Qianwen Wang, Shreyas Havaldar, Akhil Vaid, Jure Leskovec, Girish Nadkarni, Benjamin S. Glicksberg, Nils Gehlenborg, Marinka Zitnik

## Abstract

Drug repurposing – identifying new therapeutic uses for approved drugs – is often serendipitous and opportunistic, expanding the use of drugs for new diseases. The clinical utility of drug repurposing AI models remains limited because the models focus narrowly on diseases for which some drugs already exist. Here, we introduce TxGNN, a graph foundation model for zero-shot drug repurposing, identifying therapeutic candidates even for diseases with limited treatment options or no existing drugs. Trained on a medical knowledge graph, TxGNN utilizes a graph neural network and metric-learning module to rank drugs as potential indications and contraindications across 17,080 diseases. When benchmarked against eight methods, TxGNN improves prediction accuracy for indications by 49.2% and contraindications by 35.1% under stringent zero-shot evaluation. To facilitate model interpretation, TxGNN’s Explainer module offers transparent insights into multi-hop medical knowledge paths that form TxGNN’s predictive rationales. Human evaluation of TxGNN’s Explainer showed that TxGNN’s predictions and explanations perform encouragingly on multiple axes of performance beyond accuracy. Many of TxGNN’s novel predictions align with off-label prescriptions clinicians make in a large healthcare system. TxGNN’s drug repurposing predictions are accurate, consistent with off-label drug use, and can be investigated by human experts through multi-hop interpretable rationales.

## Main

There is a pressing need to develop therapies for many diseases that currently lack treatments^1, 2^. Of over 7,000 rare diseases worldwide, only 5-7% of rare diseases have FDA-approved drugs^3^. Leveraging existing therapies and expanding their use by identifying new therapeutic indications via drug repurposing can alleviate the global disease burden. Using safety and efficacy data for existing drugs, drug repurposing can expedite translation to the clinic and lower development costs than designing drugs from scratch^4^ (Figure 1a). The premise behind repurposing is that drugs can have pleiotropic effects beyond the mechanism of action of their direct targets^5^. Approximately 30% of FDA-approved drugs are issued at least one post-approval new indication, and many drugs have accrued over ten indications over the years^6^. However, most repurposed drugs are the result of serendipity^7, 8^ – either observed through off-label prescriptions written by clinicians, as with gabapentin and bupropion^8^, or discovered through patient experience, as with sildenafil^6^. Predicting the efficacy of all drugs against all diseases would enable us to select drugs with fewer side effects, design more effective treatments targeting multiple points in a disease pathway, and systematically repurpose existing drugs for new therapeutic use.

**Figure 1:**
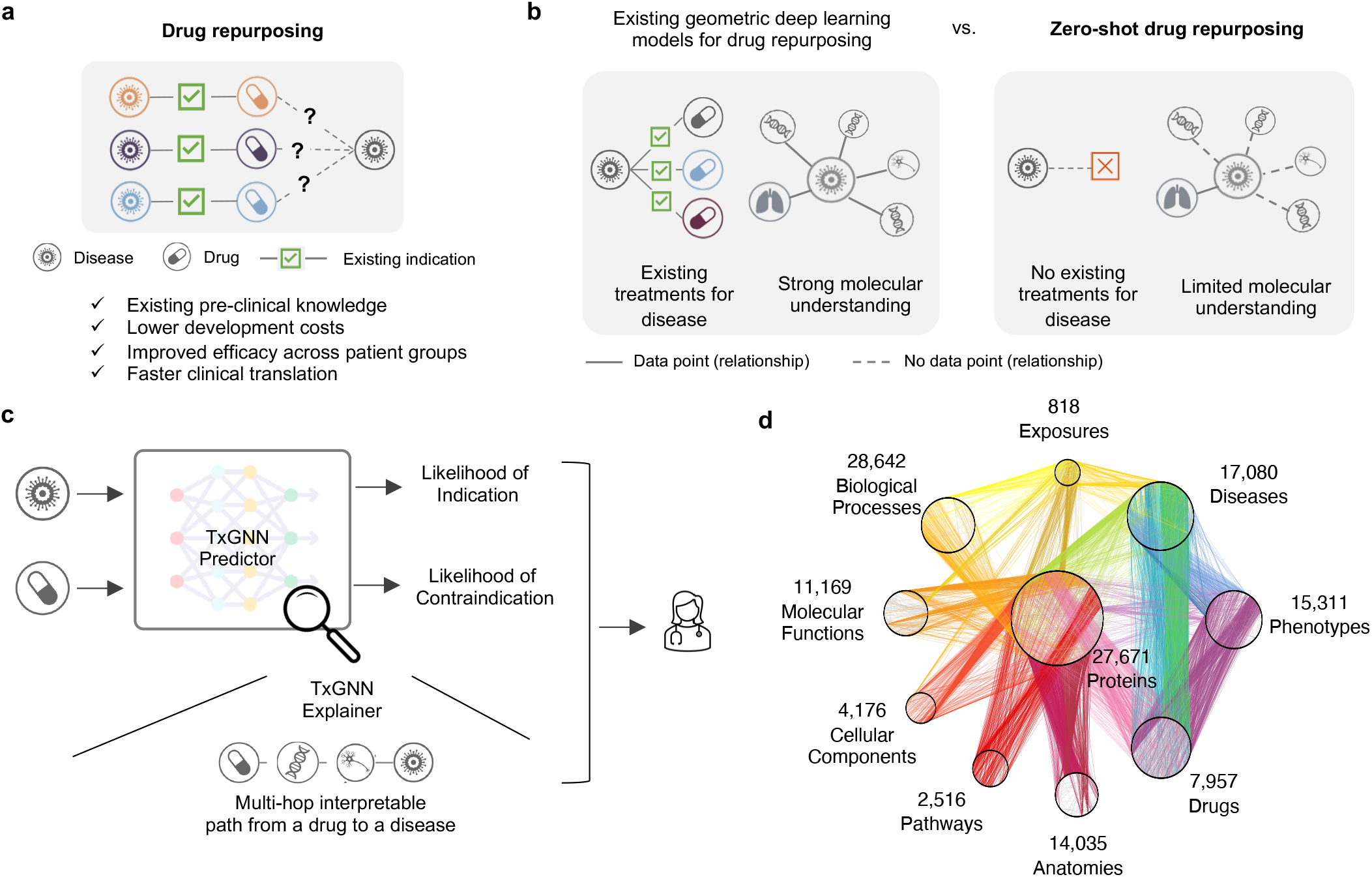
TxGNN is a graph foundation model for drug repurposing, identifying candidate drugs for diseases with limited treatment options and limited molecular data. **a**. Drug repurposing involves exploring new therapeutic applications for existing drugs to treat different diseases. Leveraging existing safety and efficacy data can dramatically cut costs and time to deliver life-saving therapeutics. **b**. Although computational drug repurposing has shown promise, it has been considered for diseases with already available treatments and well-understood molecular mechanisms. However, many diseases lack treatments and a complete understanding of disease mechanisms. These inherent constraints pose challenges for AI models. TxGNN addresses this challenge by formulating drug repurposing as a zero-shot prediction problem. **c**. TxGNN presents a novel AI framework that generates actionable predictions for zero-shot drug repurposing. TxGNN geometric deep learning model incorporates a vast and comprehensive biological knowledge graph to accurately predict the likelihood of indication or contraindication for any given disease-drug pair. Additionally, TxGNN generates explainable multi-hop paths, facilitating human understanding of how the prediction is grounded in medical knowledge. **d**. TxGNN model is trained on a medical knowledge graph of disease mechanisms across 17,080 diseases and drug mechanisms of action for 7,957 drugs.

Owing to technological advances, the effects of drugs can now be prospectively matched to new indications by systematically analyzing medical knowledge graphs^5, 9^. These strategies identify therapeutic candidates based on their impact on cell signaling, gene expression, and disease phenotypes^5, 10–12^. Machine learning has been used to analyze high-throughput molecular interactomes to unravel genetic architecture perturbed in disease^12, 13^ and help design therapies to target them^14^. To provide therapeutic predictions, geometric deep learning models optimized on large medical knowledge graphs^15^ can match disease signatures to therapeutic candidates based on networks perturbed in disease^15–18^.

Although computational approaches have identified promising repurposing candidates for complex diseases^16, 19, 20^, there remain two key challenges that could enhance the clinical relevance of repurposing predictions. First, these approaches assume that we want to make therapeutic predictions for diseases that already have existing drugs. While this is the case for some diseases^9^, a long tail of diseases does not satisfy this assumption – 92% of 17,080 diseases examined in our study have no indications. Moreover, around 95% of rare diseases have no FDA-approved drugs, and up to 85% of rare diseases do not have even one drug developed that would show promise in rare disease treatment, diagnosis, or prevention^21^. This long tail of diseases with few or no therapies and limited molecular understanding presents a challenge for drug repurposing models. Second, a repurposed indication for a therapeutic candidate can be unrelated to the indication for which the drug was initially studied. Originally developed to help with morning sickness during pregnancy, thalidomide was repurposed in 1964 for an autoimmune complication of leprosy and again in 2006 for multiple myeloma^8^. Collectively, we refer to these challenges as the zero-shot drug repurposing problem (Figure 1b). To be clinically useful, machine learning models must make “zero-shot” predictions; that is, they need to extend therapeutic predictions to diseases whose understanding is incomplete and, further, to diseases without FDA-approved drugs. Unfortunately, the ability of existing machine learning models to identify therapeutic candidates for diseases with incomplete, sparse data and zero known therapies drops drastically^16, 22^ (as we demonstrate across eight benchmarks in Figures 2c and 2d).

**Figure 2:**
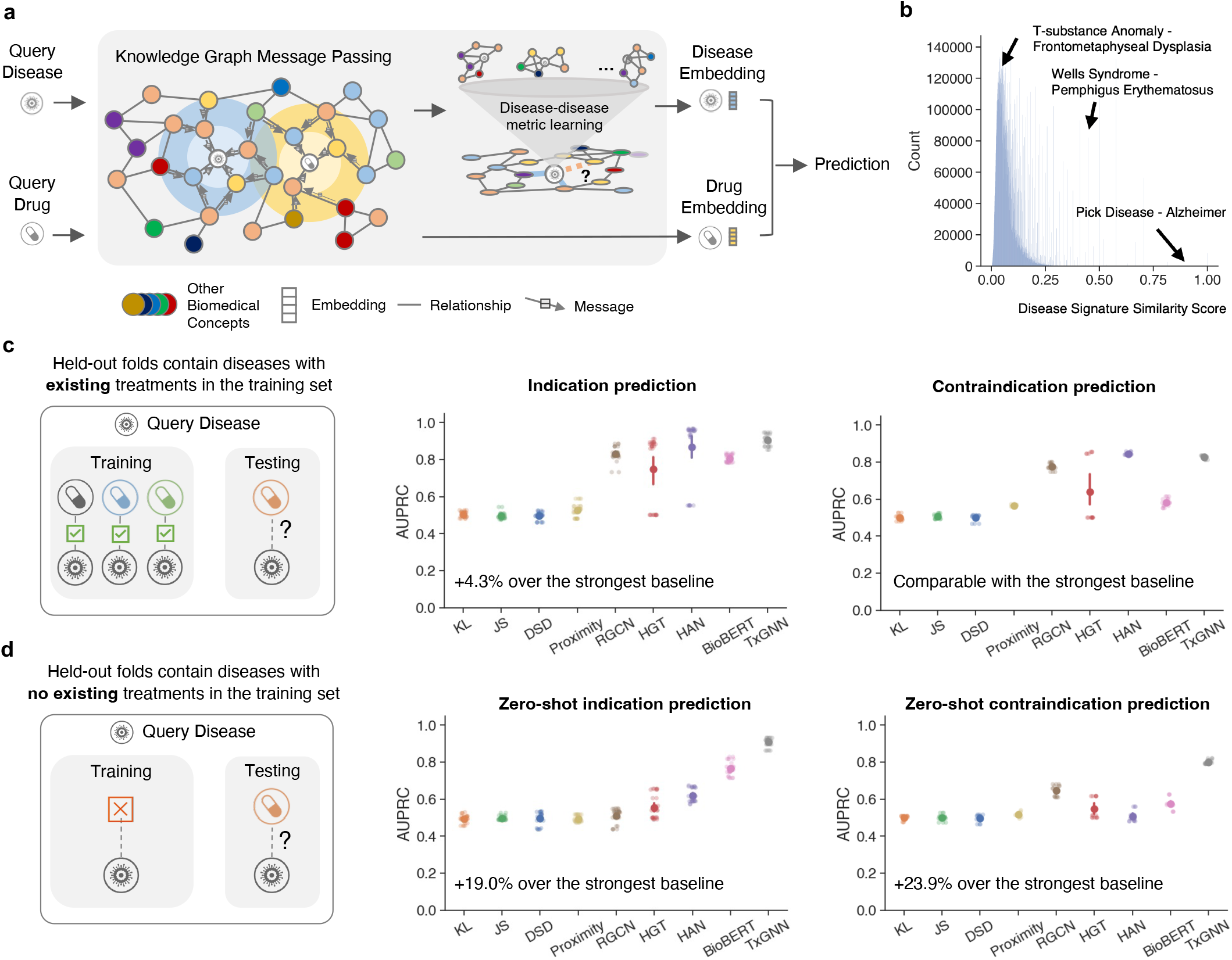
TxGNN accurately predicts drug indications and contraindications. **a**. TxGNN is a deep learning model that learns to reason over a large-scale knowledge graph to predict the relationship between drugs and disease. In zero-shot repurposing, limited indication and mechanism information is available for the query disease. Our key insight revolves around the interconnectedness of biological systems. We recognize that diseases, despite their distinctiveness, can exhibit partial similarities and share multiple underlying mechanisms. Based on this motivation, we have developed a specialized module known as disease pooling, which harnesses the power of network medicine principles. This module identifies mechanistically similar diseases and employs them to enhance the information available for the query disease. The disease pooling module has significantly improved the prioritization of repurposing candidates within zero-shot settings. **b**. The TxGNN disease similarity score provides a nuanced and meaningful measure of the relationship between diseases. This metric empowers TxGNN to discover similar diseases that can inform and enrich the understanding of query diseases lacking treatment and mechanistic information. **c**. The conventional AI-based repurposing evaluates indication predictions on diseases where the model may have seen other approved drugs during training. In this scenario, we show that TxGNN achieves good performance along with existing methods. **d**. To provide a more realistic evaluation, we introduce a novel setup for assessing zero-shot repurposing, where the model is evaluated on diseases that have no approved drugs available during training. In this challenging setting, we observe a significant degradation in performance for baseline methods. In contrast, TxGNN consistently exhibits robust performance, surpassing the best baseline by up to 19% for indications and 23.9% for contraindications. These results highlight TxGNN’s reasoning capabilities when confronted with query diseases lacking treatment options. For both **c** and **d**, the evaluation uses the area under the precision-recall curve (AUPRC) and is conducted with five random data splits (N=5). Shown is the average performance; 95% confidence intervals are represented by error bars.

Here, we introduce TxGNN, a graph foundation model for multi-disease zero-shot drug repurposing that predicts drug repurposing candidates across 17,080 diseases, including diseases without treatments (Figure 1c). Foundation models like TxGNN are transforming deep learning: instead of training disease-specific models for every disease, TxGNN is a single pre-trained model that adapts across many diseases. TxGNN is trained on a medical knowledge graph that collates decades of biological research across 17,080 diseases (Figure 1d). It uses a graph neural network model to embed drugs and diseases into a latent representation space optimized to reflect the geometry of TxGNN’s medical knowledge graph. To make zero-shot therapeutic predictions, TxGNN implements a metric learning module to transfer knowledge from treatable diseases to diseases with no treatments. Once trained, TxGNN performs zero-shot inference on new diseases without additional parameters or fine-tuning. To facilitate the interpretation of drug candidates, we develop a TxGNN Explainer module that offers transparent insights into the multi-hop interpretable paths that form TxGNN’s predictive rationales. TxGNN’s predictions and explanations are available at http://txgnn.org. Our human evaluation of TxGNN’s Explainer showed that TxGNN’s explanations perform encouragingly on multiple axes of performance, including accuracy, trust, usefulness, and time efficiency. Many of TxGNN’s novel predictions have shown alignment with off-label prescriptions made by clinicians in a large healthcare system, and TxGNN’s predictive rationales are consistent with medical reasoning.

## Results

### Overview of TxGNN zero-shot drug repurposing model

Zero-shot drug repurposing involves predicting therapeutic candidates for diseases with limited or no treatment options (Figure 1b). Mathematically, the model inputs a query drug-disease pair and outputs the likelihood of the drug acting on the disease. The gold standard labels for evaluating such a model come from our previously curated medical knowledge graph^9^ (Figure 1d, Tables S4-S5), which consists of 9,388 indications and 30,675 contraindications^23^. The medical knowledge graph covers 17,080 diseases, 92% lacking FDA-approved drugs, covering rare and less-understood complex diseases. The knowledge graph also comprises 7,957 potential drug repurposing candidates, ranging from FDA-approved drugs to experimental drugs investigated in ongoing clinical trials. Our model for zero-shot drug repurposing, TxGNN (Methods, Figure S2), operates on the principle that effective drugs directly target disease-perturbed networks or indirectly propagate therapeutic effects through disease-associated networks. TxGNN has two modules: the TxGNN Predictor module predicts drug indications and contraindications, and the TxGNN Explainer module finds interpretable multi-hop knowledge paths that connect the query drug to the query disease (Figure 1c).

The TxGNN Predictor module consists of a graph neural network (GNN) optimized on the relationships in the medical knowledge graph (Methods). Through large-scale self-supervised pre-training, the GNN produces meaningful representations for all concepts in the knowledge graph. The pre-trained model is adapted to process therapeutic tasks and predict candidate indications and contraindications of drugs across an array of diseases through fine-tuning, with no or minimal additional training of the model. TxGNN leverages an additional metric learning component for zero-shot prediction, capitalizing on the insight that diseases can share disease-associated genetic and genomic networks^10, 14^ and thus, medical knowledge of disease-associated networks can be transferred by the model from well-annotated diseases to other diseases to enhance predictions on diseases with limited treatment options (Figure 2a, Figure S1). This is achieved by creating a disease signature vector for each disease concept based on its neighbors and the topology of the local disease-associated network in the knowledge graph. The similarity between diseases is measured by the normalized dot product of their signature vectors. Since most diseases do not share underlying pathology, they have low similarity scores. In contrast, relatively high disease similarity scores (>0.2) suggest similar disease mechanisms (Figure 2b).

When querying a specific disease, TxGNN retrieves similar diseases, generates embeddings for them, and adaptively aggregates them based on their similarity to the queried disease. The aggregated output embedding summarizes knowledge borrowed from similar diseases fused with the query disease embedding. This step can be interpreted as a graph rewiring technique in the geometric machine learning literature (Figure S3). TxGNN processes different therapeutic tasks, such as indication and contraindication prediction, in a unified manner using drug and disease representations from the unified latent space of the knowledge graph (Methods). Given a query disease, TxGNN ranks drugs based on their predicted likelihood scores, offering a prioritized list of drug repurposing candidates.

While TxGNN Predictor provides likelihood scores for drug repurposing candidates, more than these are needed for trustworthy model use. Human experts seek to understand the reasoning behind these predictions to validate the model’s hypotheses and better understand candidate treatment mechanisms. To this end, TxGNN Explainer parses the knowledge graph to extract and succinctly represent relevant medical knowledge. TxGNN uses a self-explaining approach called GraphMask^24^ (Methods). GraphMask generates a sparse yet sufficient subgraph of medical concepts considered critical by TxGNN for prediction. TxGNN produces an importance score between 0 and 1 for every edge in the medical knowledge graph. It relates a drug to disease through multi-hop paths that form TxGNN’s predictive rationale, with 1 indicating the edge is vital for prediction and 0 suggesting it is irrelevant. TxGNN Explainer combines the drug-disease subgraph and edge importance scores to produce multi-hop interpretable rationales relating disease to the predicted drug. TxGNN Explainer offers granular explanations that are, as we show in a human evaluation study, aligned with human expert intuition.

We developed a human-centered tool with TxGNN’s predictions and multi-hop interpretable paths. Amongst a range of designs (Figures S4 and S5), we focused on path-based reasoning because our human evaluation study demonstrated that this design choice enhanced clinician comprehension and satisfaction^25^.

### Treatment matching and zero-shot drug repurposing

We evaluated model performance in drug repurposing across various hold-out datasets (Table S1-S2). We generated a hold-out dataset by sampling diseases from the knowledge graph. These diseases were deliberately omitted during the training phase and later served as test cases to gauge the model’s ability to generalize its insights to previously unseen diseases. These held-out diseases were chosen randomly, following a standard evaluation strategy, or specifically selected to evaluate zero-shot prediction. In our study, we used both hold-out datasets to evaluate methods. We compared TxGNN to eight methods in predicting therapeutic use. They included network medicine statistical techniques, including KL and JS divergence^16^, graph-theoretic network proximity approach^19^, diffusion state distance (DSD)^26^, state-of-the-art graph neural network methods, including relational graph convolutional networks (RGCN)^18, 27^, heterogeneous graph transformer (HGT)^28^, and heterogeneous attention networks (HAN)^29^, and a natural language processing model, BioBERT^30^ (Supplementary Note 4).

We first implemented a standard benchmarking strategy used to evaluate drug repurposing AI models, where drug-disease treatment pairs were randomly shuffled, and a subset of these pairs was set aside as a hold-out set (testing set; Figure 2c). Under this strategy, the diseases evaluated as holdouts had some drug indications and contraindications in the training dataset. Therefore, the generalization objective was to identify therapeutic candidates for diseases with some existing drugs. This evaluation method aligns with the approach predominantly used in literature^12, 14–16, 18–20^. We use the area under the precision-recall curve (AUPRC) as the evaluation metric as it measures a model’s recall and precision tradeoff at different thresholds. Our experimental results in this setting concur, with 3 of 8 existing methods achieving AUPRC greater than 0.80 and HAN as the best at 0.873 AUPRC. TxGNN also performed similarly to these established approaches. In predicting indications, TxGNN achieved a 4.3% increase in AUPRC (0.913) over HAN.

These results show that machine learning models can identify additional candidate drugs for diseases that already have some existing FDA-approved drugs. However, Duran et al.^31^ reasons that these models make predictions for a disease by retrieving drugs from the dataset that appear similar to existing treatments. This suggests the standard evaluation strategy is inappropriate for evaluating diseases without FDA-approved drugs (Figure 1b). Given this limitation, we consider models under zero-shot drug repurposing. We began by holding out a random set of diseases and then moved all their associated drugs to the hold-out set (Figure 2d). From a biological standpoint, the model was required to predict therapeutic candidates for diseases that lacked treatments, meaning it had to operate without any available data on drug similarities. In this scenario, TxGNN outperformed all existing methods by a large margin. TxGNN significantly improves over the next best method in predicting indications (19.0% AUPRC gain) and contraindications (23.9% AUPRC gain). While established methods achieved satisfactory results in conventional drug repurposing evaluations, they often fell short in challenging settings. TxGNN was the only method that achieved consistent performance across all settings.

### Zero-shot drug repurposing evaluation across disease areas

Diseases with shared mechanisms can also share effective drugs^10^. For instance, selective serotonin reuptake inhibitors (SSRIs) can address multiple psychiatric conditions, including major depressive disorder, anxiety disorder, and obsessive-compulsive disorder. If, during training, a model learns that an SSRI is indicated for major depressive disorder, it does not take a large leap to suggest that the same SSRI could be effective for obsessive-compulsive disorder during testing^22^. This phenomenon is known as shortcut learning^32^ and underlies many deep learning failures^33^. Shortcut decision rules tend to perform well on standard benchmarks but fail on challenging conditions^34^, such as predicting drugs for rare diseases or subtypes of complex diseases.

To evaluate drug repurposing models for these challenging diseases, we curated a stringent hold-out dataset that contained a group of biologically related diseases, termed disease area. For each disease area, all drug indications and contraindications were removed from the training dataset, along with a fraction of relationships between drug nodes and other medical concepts in the knowledge graph. This dataset split evaluates model performance for diseases with limited molecular data and no existing drugs (Figure 3a). Under this setup, diseases in the hold-out evaluation set have considerably fewer neighbors than in the training set (Figure S6). In this study, we considered nine diverse disease area hold-out datasets characterized in Table S3 and listed here in order of increasing disease area size: (1) diabetes-related diseases such as gestational diabetes and lipoatrophic diabetes; (2) ‘adrenal gland’ diseases, including Addison and ectopic Cushing’s syndrome; (3) ‘autoimmune’ diseases, including celiac disease and Graves’ disease; (4) ‘anemia’ with conditions such as thalassemia and hemoglobin C disease; (5) ‘neurodegenerative’ diseases include pick disease and neuroferritinopathy; (6) ‘mental health’ disorders, including anorexia nervosa and depressive disorder; (7) ‘metabolic disorder’, including macroglobulinemia and Gilbert syndrome; (8) ‘cardiovascular’ diseases, including long QT syndrome and mitral valve stenosis; (9) ‘cancerous’ diseases, including neurofibroma and Leydig cell tumors.

**Figure 3:**
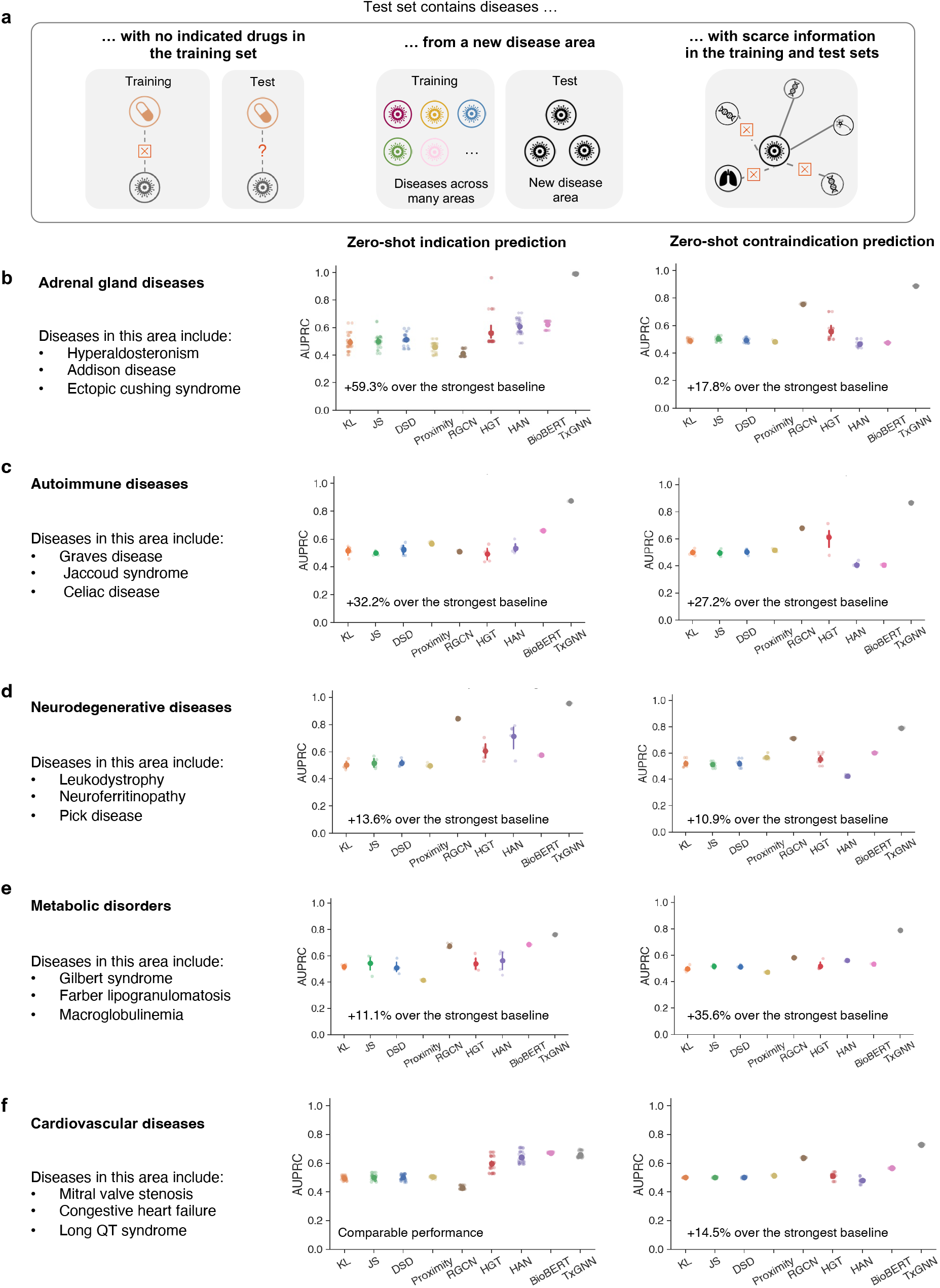
TxGNN predicts drug indications and contraindications across challenging disease areas with small molecular datasets. **a**. We construct nine ‘disease area’ splits to evaluate how well each model can generalize to new diseases when using only a limited amount of disease-associated molecular data and no information about its treatments. The diseases in the holdout set have (1) no approved drugs in training, (2) limited overlap with the training disease set because we exclude similar diseases, and (3) lack molecular data because we deliberately remove their biological neighbors from the training set. These data splits constitute challenging but realistic evaluation scenarios that mimic zero-shot drug repurposing settings. **b-f**. Holdout folds evaluate diseases related to adrenal glands, autoimmune diseases, neurodegenerative diseases, metabolic disorders, and cardiovascular diseases. Results for four disease areas, anemia, diabetes, cancer, and mental health, are provided in Figure S7. Raw scores are provided in Tables S1 and S2. TxGNN shows up to 59.3% improvement over the next best baseline in ranking therapeutic candidates, measured by AUPRC. Each method under each split is conducted with five random data splits (N=5). Shown is the average performance; 95% confidence intervals are represented by error bars.

We benchmarked TxGNN on rigorous hold-out datasets (Figure 3b-f and S7) and found consistent improvement over existing methods. TxGNN achieved relative AUPRC gains of 0.5% to 59.3% (average 25.72%) across nine disease areas for indications and 11.8% to 35.6% (average 18.67%) for contraindications. BioBERT performed best for indication prediction in 7 of 9 disease areas, while RGCN was the best baseline for contraindications in 8 of 9. However, TxGNN outperformed all methods across all nine disease areas for both tasks, demonstrating its broad generalizability and accuracy in zero-shot drug repurposing.

Visualization of TxGNN Predictor’s latent representations shows it can transfer knowledge from unrelated diseases to those with limited data (Figure S8). Evaluation metrics, including AUROC and recall, are detailed in Figures S9-S11. Ablation analyses confirmed that each component of the TxGNN Predictor is essential for its performance (Figure S12). Stress tests with additional data splits, minimal disease connections to the knowledge graph (Figure S13), masked local neighborhoods (Figure S14), and various knowledge graph configurations (Figure S15) demonstrated that TxGNN maintains strong predictive performance.

### TxGNN’s explanations reflect model’s predictive rationales

TxGNN extracts multi-hop interpretable paths as sequences of associations between medical concepts in the knowledge graph that establish a connection between a predicted drug and a predicted disease to substantiate TxGNN’s prediction. This tool isolates maximally predictive subgraphs that connect the query drug to the query disease through multiple hops, following relationships in the knowledge graph. The performance of these subgraphs is nearly equivalent to that of the entire knowledge graph. Focusing on the most predictive relationships (i.e., edges with importance scores above 0.5, representing an average of 14.9% of edges from the knowledge graph), the model’s performance showed a slight reduction from AUPRC=0.890 (STD: 0.006) to AUPRC=0.886 (STD: 0.005), indicating that TxGNN’s Explainer effectively identifies key associations^35^ and that explanations accurately reflect TxGNN’s internal reasoning. Conversely, when excluding edges deemed predictive by TxGNN and considering the remaining irrelevant relationships (i.e., edges with importance scores below 0.5, accounting for an average of 85.1% of edges), the predictive performance dropped from AUPRC=0.890 (STD: 0.006) to AUPRC=0.628 (STD: 0.026).

To assess the quality of TxGNN’s explanations, we use three established metrics^35^: insertion, which measures predictive performance using only the top K% of edges ranked highest by explanation weight; deletion, which assesses performance after removing the top K% of edges considered most explainable; stability, which evaluates the consistency of explanation weights through Pearson’s correlation before and after introducing random perturbations to the knowledge graph. We additionally experimented with three graph explainability methods: GNNExplainer^36^, Integrated Gradients^37^, and Information Bottleneck^38^. As shown in Figure S16, the top-ranked explainable edges are crucial, significantly impacting performance when either removed from or inserted into a graph. The performance remained consistent across all insertion and deletion percentages. Additionally, TxGNN Explainer demonstrated the most stable explanation weights under various levels of knowledge graph perturbation. These analyses confirm that TxGNN’s multi-hop interpretable paths capture elements of the knowledge graph that are most critical for making accurate predictions.

### TxGNN supports human-centric evaluation of drug candidates

To examine the utility of TxGNN’s multi-hop interpretable paths for human expert evaluations, we conducted a pilot human study with clinicians and scientists (see Figure S17 for the study interface). The study participants included five clinicians, five clinical researchers, and two pharmacists (7 male and 5 female experts, mean age=34.3, Figure 4c). For assessing drug-disease indication predictions, these participants were asked to evaluate 16 predictions from TxGNN, 12 of which were accurate. We recorded participants’ assessment accuracy, exploration time, and confidence scores for each prediction, totaling 192 trials (16 predictions × 12 participants, Table S4, S5). The user study took around 65 minutes on average, including assessing drug-disease indication predictions from TxGNN, a usability questionnaire, and a semi-structured interview.

**Figure 4:**
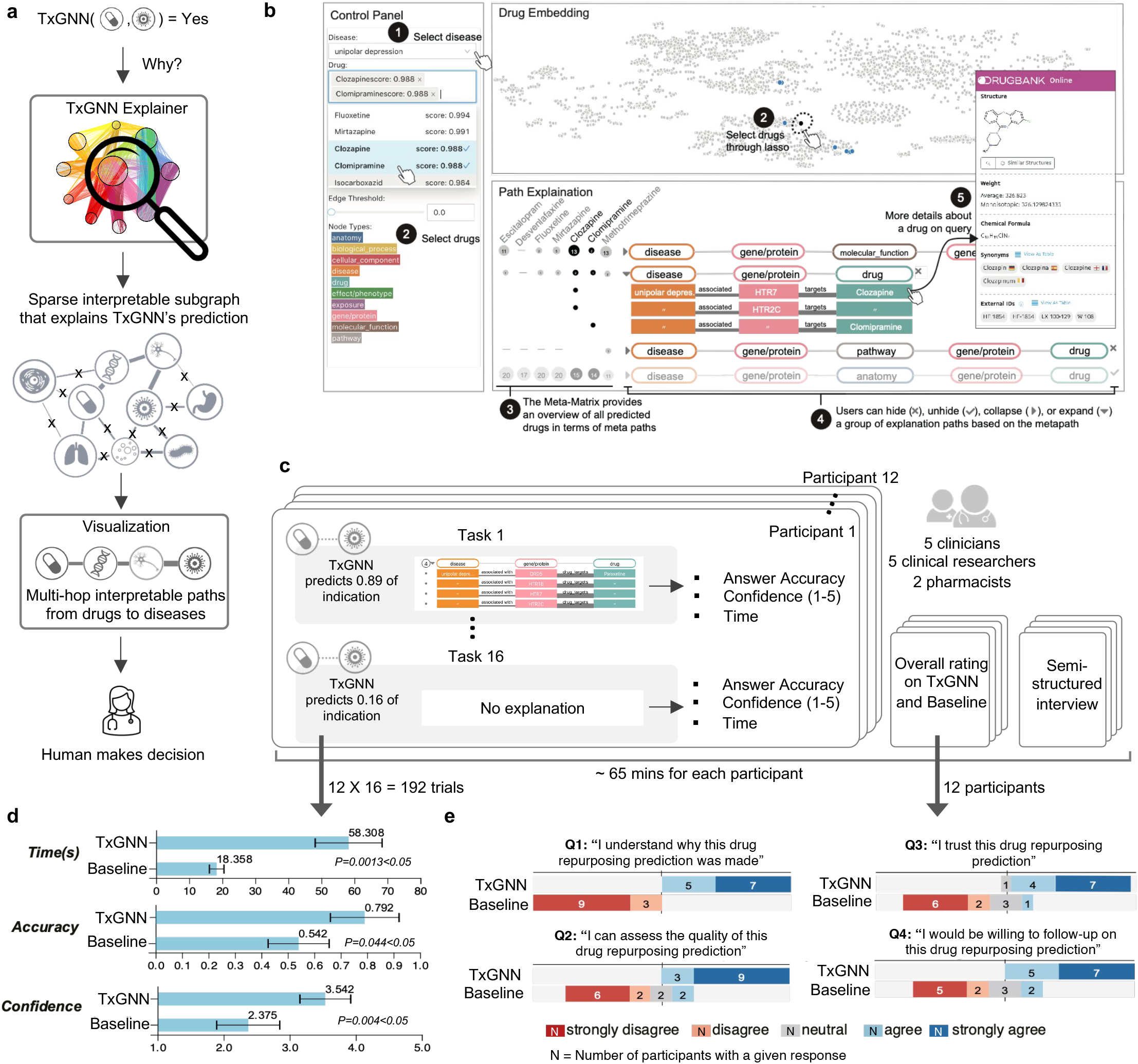
Development, visualization, and evaluation of multi-hop interpretable paths in TxGNN Explainer. **a**. Prediction alone is often insufficient for trustworthy machine learning model deployment. We develop TxGNN Explainer to aid human experts using graph AI explainability techniques. TxGNN Explainer identifies a sparse, interpretable subgraph underlying the model’s predictions. For each drug candidate, it generates a multi-hop path of biomedical concepts linking the disease to the drug. A visualization module then transforms these subgraphs into multi-hop paths that align with human cognitive processes. **b**. We design an interactive tool to help experts explore TxGNN predictions and explanations. The ‘Control Panel’ lets users select a disease and view top-ranked predictions. The ‘Edge Threshold’ module adjusts the sparsity of explanations, controlling the density of displayed multi-hop paths. The ‘Drug Embedding’ panel compares a selected drug’s position to the entire repurposing candidate library. The ‘Path Explanation’ panel shows crucial biological relations for TxGNN’s therapeutic predictions. **c**. To evaluate the usefulness of TxGNN explanations, we conducted a user study involving five clinicians, five clinical researchers, and two pharmacists. These participants were shown 16 drug-disease pairs with TxGNN’s predictions, where 12 predictions were accurate. For each pairing, participants indicated whether they agreed or disagreed with TxGNN’s predictions using the explanations provided. **d**. We compared the performance of TxGNN Explainer with a no-explanation baseline regarding user answer accuracy, task completion time, and user confidence. The results are aggregated on 192 trials (12 participants × 16 tasks) and reveal a significant improvement in accuracy (P=0.044), confidence (P=0.004), and time to think (P=0.0013) when explanations were provided. Error bars represent 95% confidence intervals, and the center of the error bar is the average performance. The statistics are computed using a two-sided Tukey’s honest significance difference test without multiple test adjustments. **e**. After the user study, participants were asked qualitative usability questions. Human experts agreed that the explanations provided by TxGNN helped assess drug repurposing candidates and instilled greater trust in the TxGNN’s predictions than using predictions alone.

In evaluating the drug repurposing candidates, participants reported a significant improvement in accuracy (+46%, *p* = 0.0443) and confidence (+49%, *p* = 0.0041) when predictions were provided with explanations. Participants took more time to think (*p* = 0.0014) to contextualize TxGNN’s explanations with their domain expertise, which led to more confident decision making (confidence +49%, *p* = 0.0041).

In the post-task questionnaires and interviews, participants reported greater satisfaction when using TxGNN Explainer compared to the baseline (Figure 4e), with 11/12 (91.6%) agreeing or strongly agreeing that the predictions and explanations provided by TxGNN were valuable. In contrast, without explanations, 8/12 (75.0%) disagreed or strongly disagreed with relying on TxGNN’s predictions. Participants expressed significantly more confidence in correct predictions made by TxGNN when the TxGNN Explainer was included (*t*(11) = 3.64, *p <* 0.01, using a two-sided Tukey’s honestly significant difference test^39^). Some participants indicated that multi-hop interpretable explanations were helpful when examining molecular target interactions identified by TxGNN Explainer and guiding evaluations of potential adverse drug events.

### Aligning TxGNN’s predictive rationales with medical evidence

We examined whether predicted drugs and their multi-hop explanations align with medical reasoning for three rare diseases. The evaluation protocol was structured into three stages (Figure 5a). Initially, a human expert queried TxGNN Predictor to identify drugs potentially repurposable for a specific disease. The TxGNN Predictor provided a candidate drug, specifying the confidence in the prediction and its comparative ranking against other candidates. Subsequently, the TxGNN Explainer was queried to elucidate why the selected drug was considered for repurposing. This model revealed its rationale through multi-hop interpretable paths linking the disease to the drug via intermediate biological interactions. In the final stage, independent medical evidence was collected and analyzed to verify the model’s predictions and explanations.

**Figure 5:**
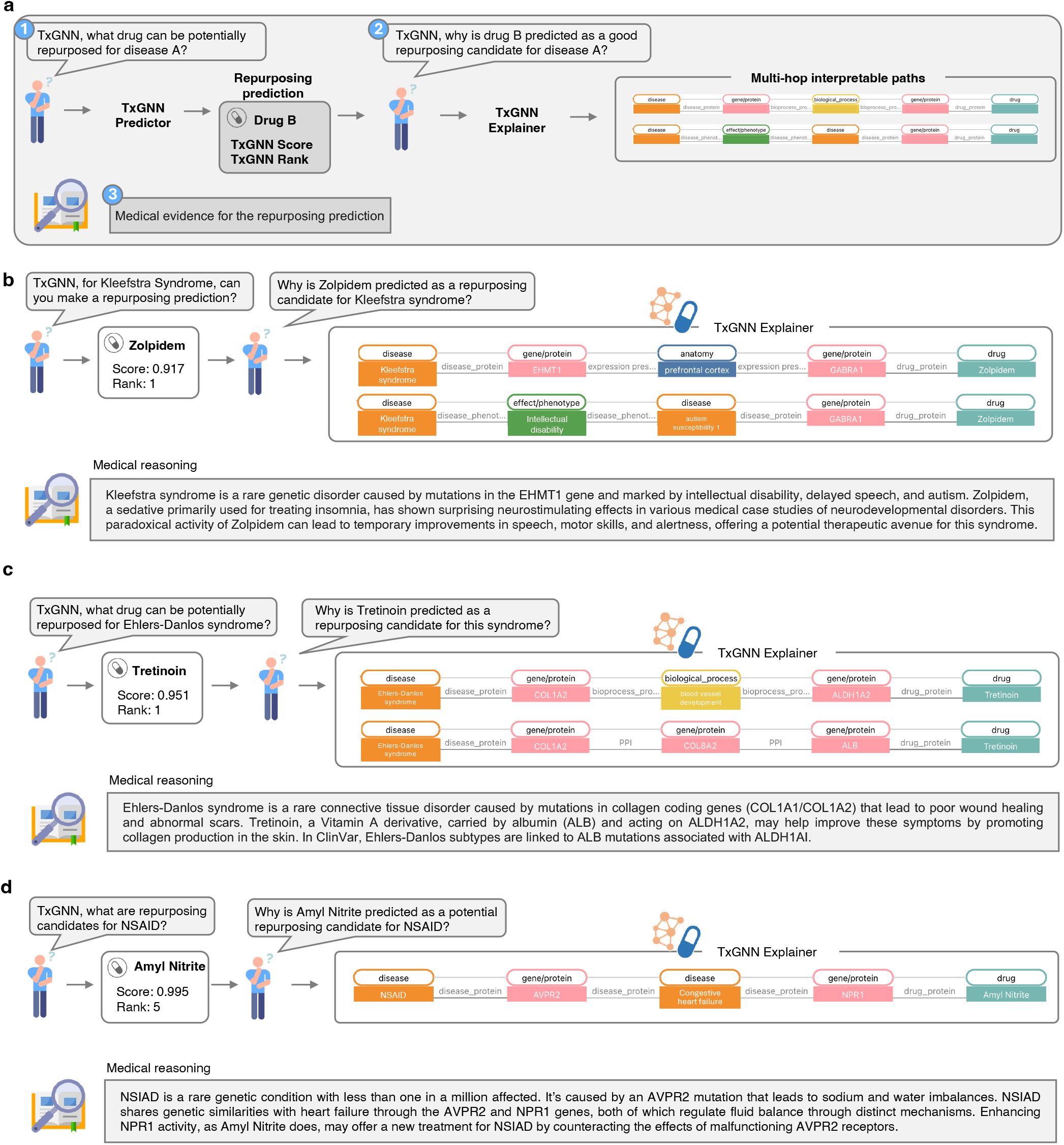
Drug repurposing predictions and multi-hop interpretable paths produced by TxGNN align with medical evidence. **a**. We assess the alignment of drug repurposing candidates identified by TxGNN with established medical reasoning across three rare diseases. The process begins with the TxGNN Predictor, which selects potential drugs for repurposing based on a disease query and continues with the TxGNN Explorer, which provides interpretable paths explaining the selection. Our case studies conclude with independent verification of the TxGNN’s predictions against clinical knowledge, showcasing the congruence between the TxGNN’s recommendations and medical insights. **b**. TxGNN predicts Zolpidem, typically used as a sedative, as a repurposing candidate for Kleefstra syndrome, characterized by developmental delays and neurological symptoms. Despite Zolpidem’s conventional inhibitory effects on the brain, TxGNN Explainer suggests its potential to enhance prefrontal cortex activity and improve cognitive functions in those with Kleefstra syndrome. TxGNN’s counterintuitive recommendation aligns with emerging clinical evidence of Zolpidem’s ability to awaken dormant neurons, potentially aiding speech, motor skills, and alertness in patients with neurodevelopmental disorders. **c**. TxGNN identifies Tretinoin as the top candidate for treating Ehlers-Danlos syndrome. TxGNN’s predictive rationale is rooted in the drug’s interactions with albumin (*ALB*) and *ALDH1A2*, which aligns with medical insights about Ehlers-Danlos syndrome regarding collagen loss and inflammation mitigation. **d**. TxGNN identifies Amyl Nitrite as a therapeutic option for the nephrogenic syndrome of inappropriate antidiuresis (NSIAD). In NSIAD, an *AVPR2* mutation leads to water and sodium imbalances. TxGNN Explorer points out the connection between NSIAD and Amyl Nitrite through congestive heart failure, a condition with similar fluid retention issues, by exploring gene interactions (*AVPR2* and *NPR1*) that regulate electrolyte balance.

First, we examined TxGNN’s predictions for Kleefstra syndrome, a rare disease caused by mutations in the *EHMT1* gene. This condition leads to speech delays, autism spectrum disorder, and childhood hypotonia, often featuring underdeveloped brains with dormant neuronal pathways. On querying the TxGNN Predictor, zolpidem was recommended as the number one drug repurposing candidate (Figure 5b). Initially, zolpidem seemed problematic for underdeveloped brains due to its sedative effect on GABA-A receptors (*GABRG2* gene). However, TxGNN Explainer indicated that zolpidem’s action on *GABRG2* might reduce autism susceptibility and improve pre-frontal cortex function. Surprisingly, zolpidem has shown stimulative effects in neurological conditions, temporarily awakening underactive neurons, suggesting a potential therapeutic use for neurodevelopmental disorders^40^. This paradoxical improvement can enhance speech, motor skills, and alertness in individuals with severe brain injuries or neurodevelopmental disorders, as supported by anecdotal evidence and some clinical studies^41, 42^. TxGNN’s prediction and explanatory rationale align with medical evidence about zolpidem’s paradoxical mechanism of action despite these clinical cases were not seen by the model during training.

Next, we examined TxGNN’s prediction of tretinoin for Ehlers-Danlos syndrome, a rare connective tissue disorder affecting 1-9 individuals per 100,000. This disorder results from mutations in collagen-coding genes (*COL1A1* and *COL1A2*) and is characterized by impaired wound healing and atypical scars. TxGNN Predictor ranks tretinoin, a vitamin A derivative used for acne, as the top drug repurposing candidate. Tretinoin, transported by albumin (*ALB*) and targeting *ALDH1A2*, helps mitigate collagen loss and inflammation, as highlighted in TxGNN’s predictive rationale (Figure 5c), indicating that TxGNN’s predictive rationale is aligned with medical reasoning. Tretinoin may help in Ehlers-Danlos syndrome by potentially enhancing wound healing and improving the appearance of scars due to its ability to stimulate collagen production in the skin. Further, some subtypes of Ehlers-Danlos syndrome have been associated with a pathogenic mutation in the *ALB* gene in Landrum et al.^43^ and weakly linked to *ALDH1AI* in Javed et al.^44^. TxGNN Explainer’s reasoning about the pathways that connect tretinoin to Ehlers-Danlos syndrome was consistent with medical evidence.

In the final example, we looked at a rare condition, nephrogenic syndrome of inappropriate antidiuresis (NSIAD). This disease is characterized by water and sodium imbalance caused by a mutation in the *AVPR2* gene. Patients with congestive heart failure face similar fluid retention challenges, and congestive heart failure has been strongly associated with both *AVPR2* and *NPR1* genes^45–47^. TxGNN Predictor identified amyl nitrite among the top-5 drugs (Figure 5d). TxGNN Explainer suggested that the relationship between NSIAD and amyl nitrite passes through *AVPR2*, congestive heart failure, and *NPR1. AVPR2* and *NPR1* genes play pivotal roles in regulating fluid and electrolyte balance via complementary but distinct pathways. *AVPR2* contributes to water retention and urine concentration, whereas *NPR1* facilitates vasodilation, lowers blood pressure, and enhances water excretion^48^. Enhancing *NPR1* activity could counteract the excessive water reabsorption caused by the malfunctioning *AVPR2* receptors in NSIAD patients. Amyl nitrite, which targets the *NPR1* gene, emerges as a potential therapeutic option for NSIAD, confirming the consistency of TxGNN’s explanations with medical evidence.

### Evaluation of TxGNN using electronic medical records

TxGNN’s strong performance suggests that its novel predictions—*i*.*e*., drugs not yet clinically approved for a disease but ranked highly by TxGNN —may hold a potential clinical value. As these therapies have not yet been approved for treatment, there is no established gold standard against which to validate them. Recognizing the longstanding clinical practice of off-label drug prescription, we used the enrichment of disease-drug pair co-occurrence in a health system’s electronic medical records (EMRs) as a proxy measure of being a potential indication. From the Mount Sinai Health System medical records, we curated a cohort of 1,272,085 adults with at least one drug prescription and one diagnosis each (Figure 6a). This cohort was 40.1 percent male, and the average age was 48.6 years (STD: 18.6 years). The demographic breakdown is in Figure 6b-c. Diseases were included if at least one patient was diagnosed with it, and drugs were included if prescribed to a minimum of ten patients (Table S6 and Methods), resulting in a dataset of 478 diseases and 1,290 drugs (Figure 6d).

**Figure 6:**
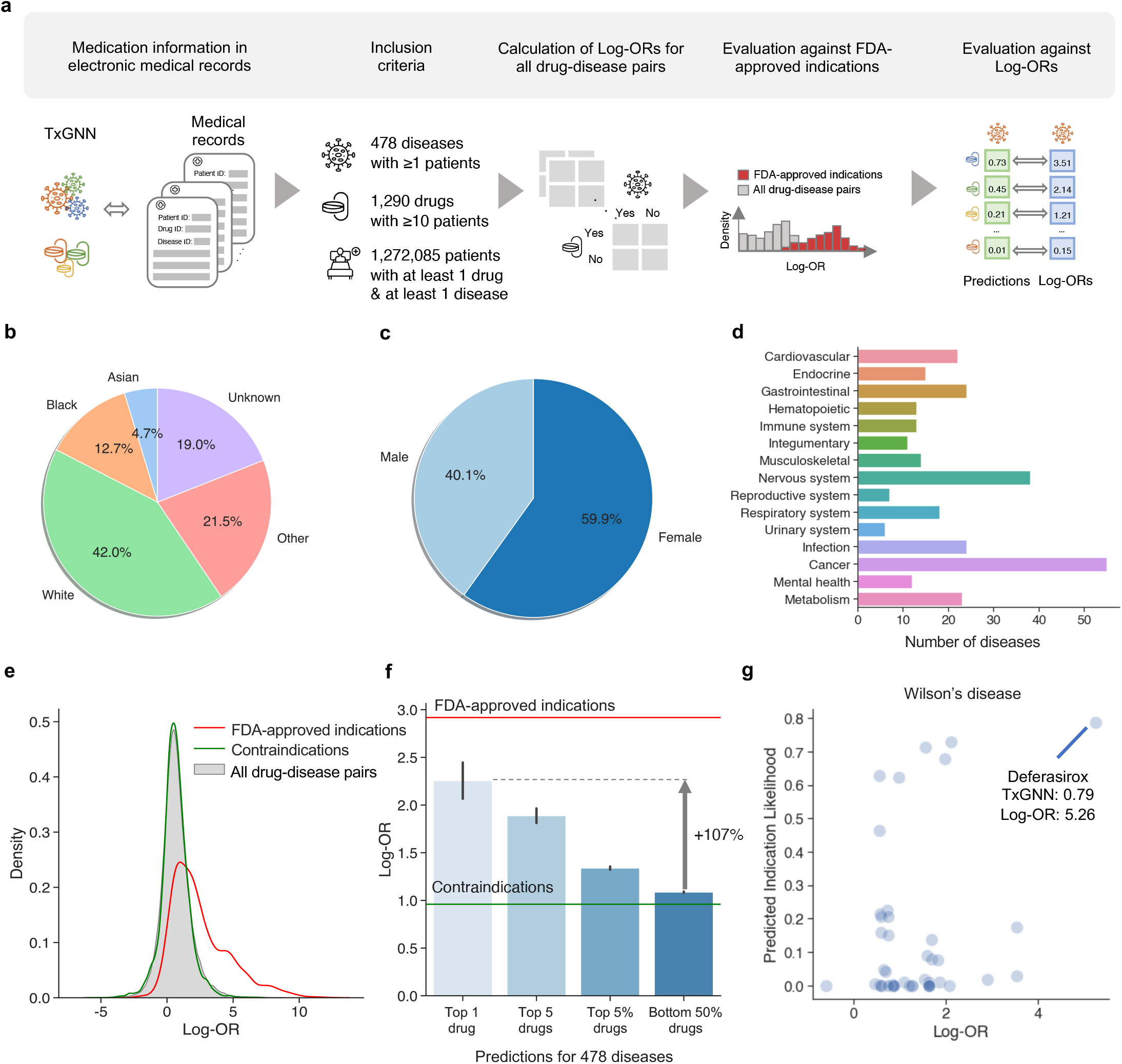
Evaluating TxGNN’s novel predictions in a large healthcare system. **a**. We illustrate the steps to evaluate TxGNN’s novel indication predictions using electronic medical records (EMRs). First, we matched the drugs and diseases in the TxGNN knowledge graph to the EMR database, resulting in a curated cohort of 1.27 million patients spanning 478 diseases and 1,290 drugs. Next, we calculated the log-odds ratio (log-OR) for each drug-disease pair to indicate drug usage for specific diseases. We validated the log-OR metric as a proxy for clinical usage by comparing drug-disease pairs against FDA-approved indications. Finally, we evaluated TxGNN’s novel predictions to determine if their log-ORs exhibited enrichment within the medical records. **b**. The racial diversity within the patient cohort. **c**. The sex distribution of the patient cohort. **d**. The medical records encompassed a diverse range of diseases spanning major disease areas. **e**. In validating log-ORs as a proxy metric for clinical prescription, we observed that while most drug-disease pairs exhibited low log-OR values, there was a significant enrichment of log-OR values for FDA-approved drugs. Additionally, we noted that contraindications displayed similar log-OR values to the general non-indicated drug-disease pairs, minimizing potential confounders such as adverse drug effects. **f**. We evaluated log-ORs for the novel indications proposed by TxGNN. The y-axis represents the log-OR of the disease-drug pairs, serving as a proxy for clinical usage. We ranked TxGNN’s predictions for each disease and extracted the average Log-OR values for the top-1 predicted drug (N = 470), top-5 predicted drugs (N = 2,314), top 5% predicted drugs (N = 27,618), and bottom-50% predicted drugs (N = 123,718). The red line represents the average log-OR for FDA-approved indications, while the green line represents the average log-OR for contraindications. Predicted drugs are consistent with off-label prescription decisions made by clinicians. The error bar is a 95% confidence interval. **g**. We provide a case study of TxGNN’s predicted scores plotted against the log-OR for Wilson’s disease. Each point on the plot represents a therapeutic candidate. The top-1 most likely drug identified by TxGNN is highlighted, indicating its associated TxGNN and log-OR scores.

Across these medical records, we measured disease-drug co-occurrence enrichment as the ratio of the odds of using a specific drug for a disease to the odds of using it for other diseases. We derived 619,200 log-odds ratios (log-ORs) for every drug-disease pair and applied necessary statistical corrections (Methods). We found that FDA-approved drug-disease pairs exhibited significantly higher log-ORs than other pairs (Figure 6e). Contraindications represented a potential confounding factor in this analysis because adverse drug events could increase the co-occurrence between drug-disease pairs. However, in our study of contraindications, we found no significant enrichment in the co-occurrence of drug-disease pairs, which suggested that adverse drug effects were not a major confounding factor.

For each of the 478 EMR-phenotyped diseases, TxGNN produced a ranked list of therapeutic candidates. We omitted drugs already linked to the disease, categorized the remaining novel candidates into top-1, top-5, top-5%, and bottom-50%, and calculated their respective mean log-ORs (Figure 6f). The top-ranked (top-1) predicted drugs had, on average, a 107% higher log-OR than the mean log-OR of the bottom-50% predictions. This suggested that TxGNN’s top candidate had much higher enrichment in the medical records and, thereby, had a greater likelihood of being a relevant indication. In addition, the log-OR increased as we broadened the fraction of retrieved candidates, suggesting that TxGNN’s prediction scores were meaningful in capturing the likelihood of indication. Although the average log-OR stands at 1.09, the top-1 therapeutic candidate predicted by TxGNN had a log-OR of 2.26, approaching the average log-OR of 2.92 for FDA-approved indications, indicating the enrichment of off-label drug prescriptions among TxGNN’s top-ranked predictions.

Examining TxGNN’s predicted drugs for Wilson’s disease, a rare disease-causing excessive copper accumulation that frequently instigates liver cirrhosis in children (Figure 6g), we observed that TxGNN predicts the likelihood close to zero for most drugs, with only a select few drugs highly likely to be indications. TxGNN ranked Deferasirox as the most promising candidate for Wilson’s disease. Wilson’s disease and Deferasirox had a log-OR of 5.26 in the medical records, and literature indicates that Deferasirox may effectively eliminate hepatic iron^49^. In a separate analysis, we evaluated TxGNN on ten recent FDA approvals introduced after the knowledge cutoff date (Table S7). TxGNN consistently ranked newly introduced drugs favorably and, in two instances, placed the newly approved medications within the top 5% of predicted drugs.

## Discussion

Drug repurposing has been embraced as a drug discovery approach to address the productivity issues of cost, time to market, and the inherent risks of developing entirely new drugs. While the conventional ‘one disease–one model’ approach has been used for drug repurposing to enhance success rates, most successful cases have resulted from unexpected findings in clinical and preclinical in vivo settings. We propose that a comprehensive approach to drug repurposing can be realized using a multi-disease predictive strategy. Existing predictive models often assume that effective drugs exist for a disease or closely related diseases. This assumption overlooks a vast array of diseases—92% of the 17,080 we analyzed—lacking pre-existing indications and known molecular target interactions. Addressing the needs of these diseases, many of which are complex, neglected, or rare, is a clinical priority^2, 50^. We define this challenge as zero-shot drug repurposing. We develop TxGNN, a graph foundation model that addresses this challenge head-on, specifically targeting diseases with limited data and treatment opportunities. TxGNN achieves state-of-the-art performance in drug repurposing by leveraging a network medicine principle focusing on disease-treatment mechanisms^15^. When asked to suggest therapeutic candidates for a disease, TxGNN identifies diseases with shared pathways, phenotypes, and pathologies, extracts relevant knowledge, and fuses it into the disease of interest. TxGNN generalizes to diseases with few treatment options by modeling latent relationships between diseases and performing zero-shot inference for diseases the model never encountered during training. The design behind TxGNN enables effective zero-shot drug repurposing and can be adapted for other use cases, such as drug target discovery and targeted therapy selection.

TxGNN is a unified model for predicting indications and contraindications across 17,080 diseases, suitable for early drug repurposing beyond single therapeutic areas. Our findings suggest that multi-disease predictive models yield more repositioned drug candidates than single-area approaches. Predicted candidates align with off-label prescription rates in electronic medical records and match with the biomedical consensus of human experts. While these estimates suggest beneficial therapeutic potential for existing drugs, predicted drugs would need extensive screening to establish safety and efficacy and determine other drug parameters, such as drug dosage and the sequence and timing of treatments.

TxGNN generates multi-hop interpretable explanations, offering rationales for predicted drugs. These rationales can be analyzed to assess if predicted drugs might elicit additional biological responses, considering the original indication or molecular target interactions identified by TxGNN. A pilot human evaluation showed that experts could more effectively examine predicted drugs and identify failure points with multi-hop explanations than alternative explanation visualizations. These findings confirm the importance of considering clinical needs and explainability when integrating machine learning models into discovery workflows^51^.

While TxGNN demonstrates promising performance for zero-shot drug repurposing, its capabilities depend on the quality of medical knowledge graphs. These graphs may need more comprehensive information on host-pathogen interactions, necessary repurposing drugs for infectious diseases, and information on the pathogenicity of genetic variants, which are crucial for identifying repurposing opportunities for genetic diseases^52^. Challenges such as data biases and potentially outdated information in the knowledge graph must be addressed. Strategies for overcoming these issues include using techniques for continual learning and model editing^53^ and data management approaches for automatically updating knowledge graphs^9^ when new data becomes available. Another fruitful future direction is using uncertainty quantification techniques to evaluate the reliability of model predictions^54^. We also envision integrating patient information with medical knowledge graphs to provide personalized drug repurposing predictions. Our pilot human evaluation engaged a small number (N=12) of clinicians and scientists and prioritized an in-depth analysis with a small but qualified group of human experts over a broader study with a larger, potentially less specialized participant pool. While the results were encouraging and this participant number is representative of related studies that evaluate highly specialized tools^55, 56^, a human evaluation study with a larger sample size could incorporate a greater diversity of user expertise and consider various drug repurposing use cases. Despite the promising performance of TxGNN’s predictions on medical records, unaccounted confounders and selection biases might have limited the ability to draw conclusions from the calculated drug enrichment scores.

TxGNN’s zero-shot drug repurposing capability allows the model to predict drugs for diseases with limited treatment options and scarce information. Multi-hop interpretable predictive rationales can enhance transparent use of TxGNN, fostering trust and aiding human experts. TxGNN streamlines drug repurposing prediction, especially when the limited availability of disease-specific datasets hinders drug development. Multi-disease models like TxGNN highlight the potential for artificial intelligence models to help with the development of new therapeutics.

## Data Availability

The TxGNN's project website is at https://zitniklab.hms.harvard.edu/projects/TxGNN. Therapeutics-centered knowledge graph is available at https://dataverse.harvard.edu/dataset.xhtml?persistentId=doi:10.7910/DVN/IXA7BM under a persistent identifier https://doi.org/10.7910/DVN/IXA7BM. We have deposited the knowledge graph and all relevant intermediate files in this repository. Python implementation of the methodology developed and used in the study is available via the project website at https://zitniklab.hms.harvard.edu/projects/TxGNN. The code to reproduce results, documentation, and usage examples are at https://github.com/mims-harvard/TxGNN. To facilitate the usage of the algorithm, we developed a TxGNN Explorer, a web-based app available at http://txgnn.org to access TxGNN's predictions.

https://zitniklab.hms.harvard.edu/projects/TxGNN

http://txgnn.org

https://github.com/mims-harvard/TxGNN

## Acknowledgements

We gratefully acknowledge the support of NIH R01-HD108794, NSF CAREER 2339524, US DoD FA8702-15-D-0001, awards from Harvard Data Science Initiative, Amazon Faculty Research, Google Research Scholar Program, AstraZeneca Research, Roche Alliance with Distinguished Scientists, Sanofi iDEA-iTECH Award, Pfizer Research, Chan Zuckerberg Initiative, John and Virginia Kaneb Fellowship award at Harvard Medical School, Biswas Computational Biology Initiative in partnership with the Milken Institute, Harvard Medical School Dean’s Innovation Awards for the Use of Artificial Intelligence, and Kempner Institute for the Study of Natural and Artificial Intelligence at Harvard University. P.C. was supported, in part, by the Harvard Summer Institute in Biomedical Informatics. Any opinions, findings, conclusions, or recommendations expressed in this material are those of the authors and do not necessarily reflect the views of the funders.

## Authors contribution

P.C. retrieved, processed, and analyzed the knowledge graph. K.H. and P.C. developed and implemented new machine learning methods, benchmarked machine learning models, and analyzed model behavior, all under the guidance of M.Z. Q.W. and N.G. implemented the clinician-centered visual explorer of model predictions and performed a user study to evaluate its usability. S.H., A.V., G.N., and B.S.G. performed a validation study examining new predictions of therapeutic use through the electronic health record system. K.H., P.C., Q.W., S.H., A.V., J.L., G.N., B.S.G., N.G., and M.Z. contributed new analytic tools and wrote the manuscript. All authors discussed the results and contributed to the final manuscript. M.Z. designed the study.

## Inclusion and ethics statement in global research

We have complied with all relevant ethical regulations. Our research team represents a diverse group of collaborators. Roles and responsibilities were clearly defined and agreed upon among collaborators before the start of the research. All researchers were included in the study design, study implementation, data ownership, intellectual property, and authorship of publications. Our research did not face severe restrictions or prohibitions in the setting of the local researchers, and no specific exceptions were granted for this study in agreement with local stakeholders. Animal welfare regulations, environmental protection, risk-related regulations, and transfer of biological materials, cultural artifacts, or associated traditional knowledge out of the country do not apply to our research. Our research does not result in stigmatization, incrimination, discrimination, or personal risk to participants. Appropriate provisions were taken to ensure the safety and well-being of all participants involved. Our team was committed to promoting equitable access to resources and benefits resulting from the research.

## Methods

Most data used for this study were obtained from publicly available knowledge repositories. For internal data, the Institutional Review Board at Mount Sinai, New York City, U.S., approved the retrospective analysis of internal electronic medical records. All internal EMRs were deidentified before computational analysis and model development. Patients were not directly involved or recruited for the study. Informed consent was waived for analyzing EMRs retrospectively.

### Curation of a medical knowledge graph dataset

The knowledge graph is heterogeneous, with ten types of nodes and 29 types of undirected edges. It contains 123,527 nodes and 8,063,026 edges. Tables S2 and S3 show a breakdown of nodes by node type and edges by edge type, respectively. The knowledge graph and all auxiliary data files are available via Harvard Dataverse at https://doi.org/10.7910/DVN/IXA7BM. Supplementary Note S1 provides detailed information about datasets and curation of the knowledge graph.

### Problem definition

We are given a heterogeneous knowledge graph (KG) *G* = (*𝒱, ℰ, 𝒯*_*ℛ*_) with nodes in the node set *i* ∈ *𝒱*, edges *e*_*i,j*_ = (*i, r, j*) in the edge set *ℰ*, where *r* ∈ *𝒯*_*ℛ*_ indicates the relation type, *i* is called the head/source node and *j* is called the tail/target node. Each node also belongs to a node type set *𝒯*_*𝒱*_. Each node also has an initial embedding, which we denote as 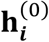. Given a disease *i* and drug *j*, we want to predict the likelihood of the drug being indicated or contraindicated for the disease. Our approach induces inductive priors by incorporating factual knowledge from the KG into the model, enhancing its reasoning capabilities for hypothesis formation and drug candidate prediction. Detailed experimental protocols, including data split curation, negative sampling, hyperparameter tuning, and additional details, are described in Supplementary Note S4.

### Overview of TxGNN approach

TxGNN is a deep learning approach for mechanistic predictions in drug discovery based on molecular networks perturbed in disease and targeted by therapeutics. TxGNN is composed of four modules: (1) a heterogeneous graph neural network-based encoder to obtain biologically meaningful network representation for each biomedical entity; (2) a disease similarity-based metric learning decoder to leverage auxiliary information to enrich the representation of diseases that lack molecular characterization; (3) an all-relation stochastic pre-training followed by a drug-disease centric full-graph fine-tuning strategy; (4) a graph explainability module to retain a sparse set of edges that are crucial for prediction as a post-training step. Next, we expand each module in detail.

### Heterogeneous graph neural network encoder

Our objective is to learn a general encoder of a biomedical knowledge graph by learning a numerical vector (embedding) for each node, encapsulating the biomedical knowledge contained within its neighboring relational structures. This involves transforming initial node embeddings using a sequence of local graph-based non-linear function transformations to refine embeddings^57^. These transformations are subject to iterative optimization, guided by a loss function that minimizes incorrect drug predictions. The system converges to an optimized set of node embeddings through this process.

#### Step 1: Initializing latent representations

We denote the input node embedding **X**_*i*_ for each node *i*, which is initialized using Xavier uniform initialization. For every layer *l* of message-passing, there are the following three stages:

#### Step 2: Propagating relation-specific neural messages

For every relation type, first calculates a transformation of node embedding from the previous layer 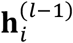, where the first layer 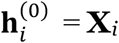. This is achieved via applying a relation-specific weight matrix 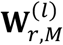 on the previous layer embedding:

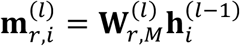

#### Step 3: Aggregating local network neighborhoods

For each node *i*, we aggregate on the incoming messages from neighboring nodes of each relation *r* denoted as 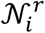 by taking the average of these messages:

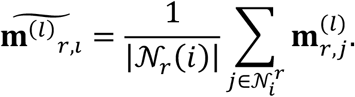

#### Step 4: Updating latent representations

We then combine the node embedding from the last layer and the aggregated messages from all relations to obtain the new node embedding:

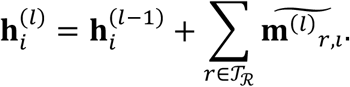

After *L* layers of propagation, we arrive at our encoded node embeddings **h**_*i*_ for each node *i*.

### Predicting drug candidates

TxGNN employs disease and drug embeddings to predict indications and contra-indications for each disease-drug pair. Considering the three relation types needing prediction, each type is assigned a trainable weight vector **w**_*r*_. The interaction likelihood for a specific relation is then determined using the DistMult approach^58^. Formally, for a disease *i*, drug *j*, and relation *r*, the predicted likelihood *p* is calculated as follows:

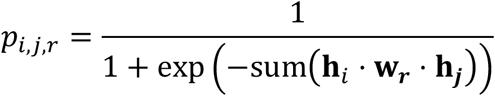

### Embedding-based disease similarity search

Research on diseases varies widely based on their prevalence and complexity. For instance, the molecular basis of many rare diseases remains poorly understood^21^. Despite this, rare diseases often offer significant opportunities for therapeutic advancements^3^. This shortage of research is evident in the biological knowledge graph, where rare diseases are characterized by a lack of relevant nodes and edges, leading to lower-quality graph embeddings. Empirical evidence indicates that GNN models exhibit substantially reduced predictive performance on splits designed to reflect the sparse nature of knowledge on these diseases, as opposed to random splits (Figure 2cd).

Network embeddings for these diseases lack significance due to sparse prior information in the KG. Thus, a model is needed to enhance these embeddings. Human physiology is an interconnected system where diseases exhibit similarities. Using a model to extract predictive information from similar but better-represented diseases in the KG, the target disease embedding can be enriched, improving model performance for the disease. To this end, TxGNN employs a three-step procedure: (1) Constructs a disease signature vector to capture complex disease similarities. (2) Uses an aggregation mechanism to combine similar disease embeddings into a comprehensive auxiliary embedding. (3) Introduces a gating mechanism to modulate the influence between the original and auxiliary disease embeddings, acknowledging that well-characterized diseases may not need supplementation. Each step is elaborated upon in the following sections.

#### Step 1: Disease signature vectors

The primary objective of this module is to derive a signature vector **p**_*i*_ for each disease *i*. Given the insufficiency of disease representations produced solely by graph neural networks, these representations are not ideal for direct similarity computations. Instead, we employ graph theoretical methods^14^ to calculate disease similarities. Additionally, variations of signature vectors are detailed in Supplementary Note S2. Specifically, we generate a vector that encapsulates the local neighborhoods surrounding a disease. For disease *i*, the signature vector is formally defined as follows:

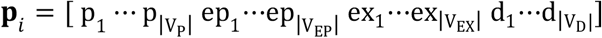

where

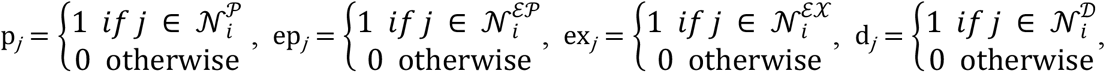

and 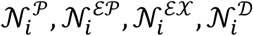 is the set of gene/protein, effect/phenotype, exposure, diseases nodes lie in the 1-hop neighborhood of disease *i*. We also adopt the dot product as the similarity measure, which means the similarity is the sum of all shared nodes across the four node types:

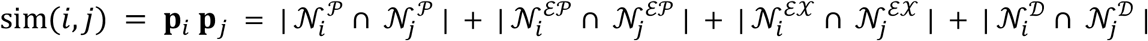

Given the signature for diseases and calculated similarities among the diseases, for a query disease, we can then obtain *k* most similar diseases for a query disease *i*:

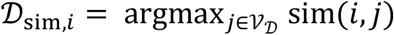

#### Step 2: Disease metric learning

Given a set of similar diseases, TxGNN generates disease embeddings that integrate various measures of disease similarity into a unified embedding, capable of augmenting the representation of a query disease that may be sparsely annotated. To achieve this, we adopt a weighted scheme, wherein each disease is weighted according to its similarity score, as follows:

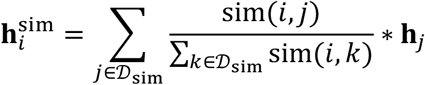

#### Step 3: Gating disease embeddings

The final stage involves updating the original disease embedding **h**_*i*_ with the disease-disease metric learning embedding 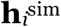 via a gating mechanism. This mechanism employs a scalar *c* ∈ [0, 1] to modulate the influence between these two embeddings. Special consideration is needed because, for well-represented diseases in the knowledge graph, the disease-disease metric learning embedding might be unnecessary and could bias the disease embedding. Conversely, this embedding can be informative for accurate prediction for diseases with no existing drugs. Using a learnable attention mechanism is ineffective, as it overvalues the original embeddings for well-represented diseases, neglecting the auxiliary embedding.

Alternatively, we introduce an approach that determines weighting based on the degree of node connectivity 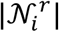 of the query drug-disease pair. A higher degree indicates the disease is better represented in the knowledge and has a denser local network neighborhood, suggesting a reduced reliance on the disease-disease metric learning embedding and vice versa. The scalar’s value is designed to be significantly high for minimal node degrees (0 or 1) and to decrease rapidly with increasing node degrees. To achieve this, we use an inflated exponential distribution density function with *λ* = 0.7:

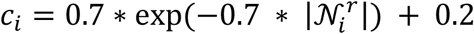

We observe the result is not sensitive to *λ* (Figure S12). Finally, we use parameter search and find optimal *λ* = 0.7. Then, we can finally obtain an augmented disease embedding:

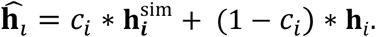

Finally, TxGNN uses augmented disease embeddings as input to the latent decoder to produce drug predictions.

### Training TxGNN deep graph models

The objective of the training process is to predict the presence of a relation between two entities within a knowledge graph. The dataset for positive samples, denoted as *𝒟*_+_, comprises all pairs (*i, j*) across various relation types *r*, with the label *y*_*i,r,j*_ = 1 indicating the presence of a relation. To generate the dataset for negative samples, *𝒟*_−_, we use a sampling technique detailed in Supplementary Note S4.3, creating counterparts for each positive pair. For a given pair *i, j* and relation type *r*, the model estimates the probability *p*_*i,r,j*_ of a relation’s existence. The training loss is then calculated using the binary cross-entropy loss formula:

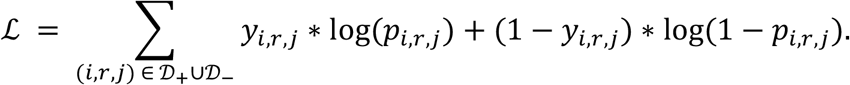

Previous research has emphasized knowledge graph completion, optimizing models across the entire spectrum of relations within a knowledge graph^59^. This approach, however, may dilute the model’s capacity to capture specific knowledge, particularly when the interest lies solely in drug-disease relations. Given that drug-disease interactions are governed by complex biological mechanisms, the extensive range of biomedical relations in a knowledge graph can offer a comprehensive view of biological systems. The primary challenge lies in optimizing performance on a select group of relations while beneficially leveraging the broader set of relations for knowledge transfer, avoiding catastrophic forgetting of general knowledge.

To address this challenge, TxGNN uses a pre-training strategy. Initially, TxGNN predicts relations across the entire KG using stochastic mini batching, encapsulating biomedical knowledge in enriched node embeddings. In the fine-tuning phase, TxGNN focuses on drug-disease relations, sharpening its ability to generate specific embeddings and optimizing drug repurposing predictions.

### Pre-training TxGNN model

TxGNN undergoes pre-training on millions of biomedical entity pairs across all relations. Due to the extensive number of edges, stochastic mini batching is used to train on subsets of pairs at each step, ensuring coverage of all data pairs within each epoch. During this phase, degree-adjusted disease augmentation is deactivated, and all relation types are treated equally. The pre-trained encoder weights are then used to initialize the encoder model for fine-tuning. It is important to note that the weights in the decoder, specifically for DistMult, **w**_*r*_, are reinitialized before fine-tuning to mitigate the risk of negative knowledge transfer.

### Fine-tuning TxGNN model

After pre-training, the model initialization encapsulates a broad spectrum of biological knowledge. The next phase refines drug-disease relation predictions by focusing solely on drug-disease pairs. Other relation types remain in the knowledge graph to facilitate indirect information flow. During fine-tuning, the model activates the degree-adjusted inter-disease embedding feature. TxGNN undergoes both pre-training and fine-tuning end-to-end. The variant with the highest validation performance is selected for test set evaluation and downstream analyses.

### Generating multi-hop interpretable explanations

In a trained drug repurposing prediction model, consider a target node *j* and a neighboring source node *i* connected by an edge *e*_*i,j*_ at layer *l*. For each relation *r*, intermediate messages 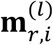 and 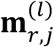 are computed. These embeddings are concatenated and input into a relation-specific, single-layer neural network parameterized by 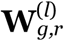. This network predicts the probability of masking the message from source node *i* during the computation of the target node *j*’s embedding. The output is processed through a gate, which includes a sigmoid layer to constrain the probability to the range [0, 1], followed by an indicator function that determines whether the edge should be dropped:

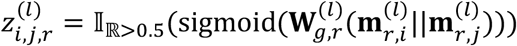

such that 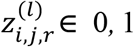. In practice, a location bias of 3 is added to the sigmoid function during initialization to ensure that its outputs are initially close to 1. This means that at the start, the gates remain open, allowing the model to adaptively close the gates and mask edges within the subgraph as needed. This approach is essential because starting with random initialization, which drops edges randomly, creates a significant discrepancy between the original and updated predictions. Consequently, the model’s primary focus shifts towards minimizing this discrepancy rather than balancing the two objectives. To refine this mechanism, when a gate outputs 0, the corresponding message is not simply removed. Instead, it is substituted with a learnable baseline vector 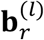 for each relation *r* and layer *l*. Therefore, the revised message from source node *i* to target node *j* is represented as follows:

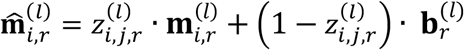

Two objectives guide the optimization of the GraphMask gate weights. The first, faithfulness, aims to ensure that the updated predictions, after applying the mask, align closely with the initial prediction outcomes. The second objective encourages the model to apply as extensive a masking as feasible. These objectives inherently entail a trade-off: increasing the extent of masking tends to enlarge the discrepancy between the updated and original predictions. This scenario is addressed through constrained optimization, employing Lagrange relaxation to balance the objectives. Specifically, the optimization seeks to maximize the Lagrange multiplier *λ* to enforce the constraint while minimizing the primary objective. The loss function employed for this purpose is formulated as follows:

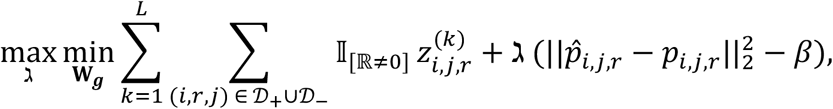

where *β* is the margin between the updated and original prediction. After the training process is complete, edges (*i, j, r*) for which 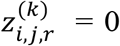 can be removed. The remaining edges serve as explanations for the model’s predictions. Additionally, the value computed prior to applying the indicator function can be employed to quantify each edge’s contribution to the prediction. This facilitates the adjustment of granular differences in the contributions. More detailed adaptations of the GraphMask approach are discussed in Supplementary Note S3.

### Pilot usability evaluation of TxGNN with medical experts

The TxGNN Explorer was developed following a user-centric design study process, as outlined in our pilot study^25^. This process involved comparing three visual presentations of GNN explanations from the user’s perspective. The findings from this comparison motivated the adoption of path-based explanations, which were preferred based on user feedback. The usability of the TxGNN Explorer was assessed through a comparison with a baseline that only displayed drug predictions and their associated confidence scores.

For this usability study, twelve medical experts (7 male and 5 female experts, average age 34.25, referred to as P1-12) were recruited through personal contacts, Slack channels, and email lists from collaborating institutions, with all participants providing informed consent. The group comprised five clinical researchers (P1-3, P11-12) and five practicing physicians (P4, P7-10), all holding

M.D. degrees, and two medical school students with prior experience as pharmacists (P5, P6). Each participant had at least five years of experience in various medical specialties.

The study was conducted remotely via Zoom in compliance with COVID-19-related restrictions. Participants accessed the study system (as shown in Figure S17) using their own computers and shared their screens with the interviewer. The sequence in which predictions were presented, along with the conditions (TxGNN Explorer or the baseline approach), was randomized and counterbalanced across participants and tasks.

In the drug assessment tasks, participants’ accuracy, confidence levels, and task completion times were evaluated across 192 trials (16 tasks × 12 participants). Specifically, participants were tasked with 1) determining the correctness of a drug prediction (i.e., if the drug could potentially be used to treat the disease) and 2) rating their confidence in their decision on a 5-point Likert scale (1=not confident at all, 5=completely confident). The system automatically logged the time taken to evaluate each prediction.

Upon completing all predictions, participants provided subjective ratings for both tasks regarding *Trust, Helpfulness, Understandability*, and *Willingness to Use*, using a 5-point Likert scale (1=strongly disagree, 5=strongly agree). Subsequent semi-structured interviews yielded insights and feedback on the tool’s predictions, explanations, and overall user experience. Each session of the user study lasted approximately 65 minutes.

### Analysis of medical records from a large healthcare system

Patient data from the Mount Sinai Health System’s electronic medical records (EMRs) in New York City, U.S., were utilized to examine patterns from predictions in clinical practice. The Mount Sinai Institutional Review Board approved the study, ensuring all clinical data were de-identified. The initial cohort included over 10 million patients, refined to those over 18 years of age with at least one drug and one diagnosis on record, resulting in 1,272,085 patients. This refined cohort comprised 40.1% males, with an average age of 48.6 years (SD: 18.6 years). The racial composition of the dataset is detailed in Table S6.

Disease and medication data were structured according to the Observational Medical Outcomes Partnership (OMOP) standard data model^60^. Predictions were generated for 1,363 diseases, identified by training a knowledge graph on 5% of randomly selected drug-disease pairs, serving as a validation set for early stopping. Disease names in the prediction dataset were aligned with SNOMED or ICD-10 codes and then mapped to OMOP concepts within the Mount Sinai data system. The analysis was restricted to diseases diagnosed in at least one patient, narrowing the focus to 478 conditions. Similarly, medication names were matched to DrugBank IDs, then to RxNorm IDs and OMOP concepts, limiting the scope to medications prescribed to at least ten patients, resulting in 1,290 medications. Drug-disease pairs were further refined to those with at least one recorded instance of a patient being prescribed the drug for the disease, leading to 1,272,085 patients. Contingency tables were created for each drug-disease pair, and the Fisher exact function was employed to calculate 2-sided odds ratios and p-values for each pair. A two-sided Bonferroni correction was applied to the p-values using the statsmodels Python library’s multi-test function, identifying statistically significant drug-disease pairs as those with p < 0.005.

## Data availability

The knowledge graph dataset is available at [Harvard Dataverse](https://dataverse.harvard.edu/dataset.xhtml?persistentId=doi%3A10.7910/DVN/IXA7BM) under a persistent identifier https://doi.org/10.7910/DVN/IXA7BM. All clinical and medical record datasets were de-identified, and dataset summary statistics were analyzed in the study. The anonymized patient data used retrospectively for this project, with institutional permission, are not publicly available due to restrictions. All requests from institution-affiliated researchers for access to processed data for purposes of study validation will be considered by the CBIPM steering committee and should be directed to Girish Nadkarni, MD, MPH, which will be handled within one month. Further information and related materials are available on TxGNN’s website (https://zitniklab.hms.harvard.edu/projects/TxGNN).

## Code availability

Python implementation of the methodology developed and used in the study is available via the project website at https://zitniklab.hms.harvard.edu/projects/TxGNN. The code to reproduce results, documentation, and usage examples is at https://github.com/mims-harvard/TxGNN. We developed a web-based app available at http://txgnn.org to access TxGNN’s predictions and predictive rationales.

