## Supplementary Materials for "A foundation model for clinician-centered drug repurposing"

### Contents


S1 Note 1: Knowledge graph data resources S3

S2 Note 2: Variations of Disease Signature Functions S4

S3 Note 3: Further details about TxGNN explainer S6

S4 Notes 4: Experimental setup and implementation details S6


### List of Figures


S1 Distribution of the number of relationships incident to diseases in the medical knowledge graph (KG). . . . . . S10
S2 Overview of TxGNN Predictor . . . . . . S11
S3 Illustration of TxGNN graph rewiring perspective. . . . . . S12
S4 Overview of TxGNN Explainer. . . . . . S13
S5 Three types of visual explanations for TxGNN predictions. . . . . . S14
S6 Number of first-hop neighbors for testing and training set diseases in the disease area splits. . . . . . S15
S7 Additional benchmarking results of TxGNN. . . . . . S16
S8 Visualizing TxGNN's latent representations. . . . . . S17
S9 Benchmarking indication predictions using Recall@100 performance metric. . . S18
S10 Benchmarking indication predictions using AUROC performance metric. . . . . S19
S11 Benchmarking contraindication predictions using AUROC performance metric. S20
S12 Ablation analyses. . . . . . S21
S13 Additional evaluation of TxGNN in settings with extremely scarce disease data. S22
S14 Robustness of TxGNN to scarce disease information. . . . . . S23
S15 Robustness of TxGNN to incomplete and noisy medical knowledge graph (KG). . S24
S16 Benchmarking explainability (XAI) algorithms and TxGNN Explainer. . . . . . S25
S17 Interface used in the usability study of TxGNN. . . . . . S26
S18 Additional benchmarking of TxGNN under varying numbers of negative samples. S27
S19 Robustness of disease signature vectors in TxGNN. . . . . . S28


### List of Tables


S1 Benchmarking models for predicting drug indications. . . . . . S29
S2 Benchmarking models for predicting drug contraindications. . . . . . S29

S3 **Statistics on disease-area-based dataset splits used to evaluate zero-shot drug repurposing.** . . . . . . . . S30
S4 **Accuracy, confidence, and time reported for participants in the human evaluation study of TxGNN.** . . . . . . . . S31
S5 **Accuracy, confidence, and time for every task in the human evaluation study of TxGNN.** . . . . . . . . S32
S6 **Demographics of the electronic health record (EHR) dataset at Mount Sinai Health System in New York City.** . . . . . . . . S32
S7 **Evaluation of TxGNN on recent FDA-approvals.** . . . . . . . . S33
S8 **Statistics on nodes in the medical knowledge graph (KG).** . . . . . . . . S34
S9 **Statistics on edges in the medical knowledge graph (KG).** . . . . . . . . S35

### Supplementary Notes

#### S1 Note 1: Knowledge graph data resources

##### S1.1 Primary data resources

The knowledge graph (KG) is compiled from many primary knowledge bases that cover ten types of biomedical entities and provide broad coverage of human disease, already-available drugs, and novel drugs in development. We briefly overview biological information retrieved from the knowledge bases, with details provided in Chandak *et al.*[1]: **Bgee**[2] contains gene expression patterns and contributes to anatomy-protein associations where gene expression was present or absent to the KG. The **Comparative Toxicogenomics Database**[3] contributes relationships between environmental exposures and proteins, diseases, other exposures, biological processes, molecular functions, and cellular components to the dataset. **DisGeNET**[4] is an expert-curated resource about the relationships between genes and human disease and provides associations of genes with diseases and phenotypes in the KG. **DrugBank**[5] is a resource that contains pharmaceutical knowledge and supplies drug-drug and drug-protein interactions to the dataset. **Drug Central**[6] curates information about 26,698 indication edges, 8,642 contraindication edges, and 1,917 off-label use edges to the KG. **Entrez Gene**[7] is a resource maintained by the NCBI that contains associations of genes with biological processes, molecular functions, and cellular components. The **Gene Ontology**[8] network describes hierarchical associations between biological processes, molecular functions, and cellular components in the KG. The **Human Phenotype Ontology**[9] provides information on disease-phenotype, protein-phenotype, and phenotype-phenotype edges in the KG. Since the **Mondo Disease Ontology**[10] harmonizes diseases from a wide range of ontologies, including OMIM, SNOMED CT, ICD, and MedDRA, it was our preferred ontology for defining diseases and also provided hierarchical disease relations. **Protein-protein interactions** are composed of experimentally-verified interactions between proteins gathered from various resources[11–18]. **Reactome**[19] is an open-source, curated database for pathways that provides pathway-pathway and protein-pathway edges to the KG. The **Side Effect Resource** (SIDER)[20] contains data about adverse drug reactions and contributes drug phenotype associations to the KG. **UBERON**[21] helps include human anatomy information in the KG.

##### S1.2 Harmonizing knowledge graph from primary data resources

To construct the knowledge graph, we harmonized ontologies for each node type, ensuring consistency by standardizing primary data sources and rectifying overlaps as described in Chandak *et al.*[1] Primary data were mapped into standardized ontologies, with drugs and diseases respectively encoded using DrugBank and Mondo Disease Ontology. For enhanced visualization in TxGNN Explainer and clarity in user studies, we introduced a 'display_relation' field as a more descriptive version of the 'relation' field, (e.g. 'disease_phenotype_negative' became 'phenotype absent').

We merged the harmonized datasets into a heterogeneous knowledge graph and extracted its largest connected component using the approach outlined in Chandak *et al.*[1]. Since the knowledge graph is designed for therapeutic use prediction, we wanted to ensure that disease nodes in the graph were meaningful representations of diseases. To this end, we adopted an approach previously validated[1] by collapsing disease nodes with nearly identical names into a single disease node. Intial disease groups were identified using automated string matching across disease names. These disease groupings were tightened using ClinicalBERT[22] embedding similarities between disease names with an empirically chosen similarity cutoff $\geq 0.98$. Finally, we manually approved the suggested disease matches and assigned names to the new groups. After grouping, 22,205 diseases in the Mondo Disease Ontology were collapsed into 17,080 grouped diseases. Node statistics and edge statistics are provided in Table S8 and S9 respectively.

#### S2 Note 2: Variations of Disease Signature Functions

We consider the following three signature functions:

- **Protein signatures (PS):** The mechanism of actions for drugs is to act upon protein targets in the disease pathway[23]. Thus, the ideal disease signature should preserve similarity in the protein target space. If two diseases have similar proteins in their corresponding disease pathways, they are more likely to have a similar treatment mechanism[11, 24]. This key observation motivates the protein signature[25]. We have a bit vector for each disease where each bit corresponds to a specific protein. A bit is flipped to one if the bit corresponds to a protein in the disease pathway. Formally, for disease $i$, the protein signature is defined as:

$$\mathbf{p}_i^{\text{PS}} = \begin{bmatrix} \text{p}_1 & \cdots & \text{p}_{|\mathcal{V}_\text{P}|} \end{bmatrix}, \tag{1}$$

  where

$$\text{p}_j = \begin{cases} 1 & \text{if } j \in \mathcal{N}_i^\text{P} \\ 0 & \text{otherwise,} \end{cases} \tag{2}$$

  and $\mathcal{N}_i^\text{P}$ is the set of proteins that lie in the 1-hop neighborhood of disease $i$ and $|\mathcal{V}_\text{P}|$ the number of total available proteins. To calculate similarity between two diseases $i, j$, we use dot product:

$$\text{sim}^{\text{PS}}i, j = \mathbf{p}_i^{\text{PS}} \cdot \mathbf{p}_j^{\text{PS}} = |\mathcal{N}_i^\text{P} \cap \mathcal{N}_j^\text{P}|. \tag{3}$$

  The similarity directly measures the number of intersecting proteins in the disease pathway of $i, j$. If the similarity is high, we know these two diseases have a larger number of intersecting diseases, which increases the probability of similar treatment mechanisms.

- **All-node-types signatures (AT):** Human knowledge about disease pathways are vastly incomplete. Thus, some diseases may not have complete protein pathways in the knowledge graph, which leads to biased protein signatures. Additional biological knowledge about diseases could potentially benefit. In the knowledge graph, other node types connect to diseases, including effect/phenotype, exposure, and disease. Since the local neighborhood can define some characteristics of diseases, we can extend the principle of protein signature, such that if two diseases share the same nodes in these additional node types, they have similar biological underpinnings. We call these all-node-types signatures. Formally, for disease $i$, the protein signature is defined as:

$$\mathbf{p}_i^{\mathrm{AT}} = \ \mathrm{p}_1 \cdots \mathrm{p}_{|\mathcal{V}_{\mathrm{P}}} \ \mathrm{ep}_1 \cdots \mathrm{ep}_{|\mathcal{V}_{\mathrm{EP}}|} \ \mathrm{ex}_1 \cdots \mathrm{ex}_{|\mathcal{V}_{\mathrm{EX}}|} \ \mathrm{ep}_1 \cdots \mathrm{ep}_{|\mathcal{V}_{\mathrm{EP}}|} \ \mathrm{d}_1 \cdots \mathrm{d}_{|\mathcal{V}_{\mathrm{D}}|} \ , \tag{4}$$

where

$$\mathrm{p}_j = \begin{cases} 1 & \text{if } j \in \mathcal{N}_i^{\mathrm{P}} \\ 0 & \text{otherwise} \end{cases}, \mathrm{ep}_j = \begin{cases} 1 & \text{if } j \in \mathcal{N}_i^{\mathrm{EP}} \\ 0 & \text{otherwise} \end{cases}, \mathrm{ex}_j = \begin{cases} 1 & \text{if } j \in \mathcal{N}_i^{\mathrm{EX}} \\ 0 & \text{otherwise} \end{cases} \mathrm{d}_j = \begin{cases} 1 & \text{if } j \in \mathcal{N}_i^{\mathrm{D}} \\ 0 & \text{otherwise} \end{cases} \tag{5}$$

and $\mathcal{N}_i^{\mathrm{EP}}, \mathcal{N}_i^{\mathrm{EX}}, \mathcal{N}_i^{\mathrm{D}}$ is the set of effect/phenotype, exposure, diseases nodes lie in the 1-hop neighborhood of disease $i$ and $|\mathcal{V}_{\mathrm{EP}}|, |\mathcal{V}_{\mathrm{EX}}|, |\mathcal{V}_{\mathrm{D}}|$ the number of total available effect/phenotype, exposure, diseases respectively. We also adopt the dot product as the similarity measure, which means the similarity is the sum of all shared nodes across the four node types:

$$\mathrm{sim}^{\mathrm{AT}} i, j = \mathbf{p}_i^{\mathrm{AT}} \cdot \mathbf{p}_j^{\mathrm{AT}} = |\mathcal{N}_i^{\mathrm{P}} \cap \mathcal{N}_j^{\mathrm{P}}| \ \ |\mathcal{N}_i^{\mathrm{EP}} \cap \mathcal{N}_j^{\mathrm{EP}}| \ \ |\mathcal{N}_i^{\mathrm{EX}} \cap \mathcal{N}_j^{\mathrm{EX}}| \ \ |\mathcal{N}_i^{\mathrm{D}} \cap \mathcal{N}_j^{\mathrm{D}}|. \tag{6}$$

- **Diffusion signatures (DS):** The above two signatures rely on the first-hop neighbor of the diseases, while higher-hop neighbors may contain useful molecular characterization. Diffusion signature simulates many random walks, where each random walk is a path of length $h$ starting from the disease $i$: $w = v_i \xrightarrow{e_{i,1}} v_1 \cdots v_{h-1} \xrightarrow{e_{h-1,h}} v_h$[26]. The set of visited nodes in the $k$-th random walk from disease node $i$ is denoted as $\mathcal{W}_i^k$. $\cap_k \mathcal{W}_i^k$ represents the total set of visited nodes across all walks, and we can calculate the normalized visitation probability for visited node $j$ as:

$$f_j = \frac{{}_k \ 1_{\mathcal{W}_i^k = j}}{{}_k \ |\mathcal{W}_i^k|} \tag{7}$$

These nodes correspond to a multi-hop snapshot of molecular interactions centering around the diseases, and the visitation probability corresponds to the influence level. Given this probability score, we can obtain the diffusion signature for disease node $i$:

$$\mathbf{p}_i^{\mathrm{DS}} = \ f_1 \ \cdots \ f_{|\mathcal{V}_{\mathrm{P}}|} \ . \tag{8}$$

For diffusion signature, we still use the dot product:

$$\begin{aligned} \mathrm{sim}^{\mathrm{DS}} i, j = \mathbf{p}_i^{\mathrm{DS}} \cdot \mathbf{p}_j^{\mathrm{DS}} &= {}_u^{|\mathcal{V}_{\mathrm{P}}|} \frac{\left({}_k \ 1_{\mathcal{W}_i^k = u}\right) \cdot \left({}_k \ 1_{\mathcal{W}_j^k = u}\right)}{{}_k \ |\mathcal{W}_i^k|^2} \\ &\sim {}_u^{|\mathcal{V}_{\mathrm{P}}|} \left({}_k \ 1_{\mathcal{W}_i^k = u}\right) \cdot \left({}_k \ 1_{\mathcal{W}_j^k = u}\right). \end{aligned} \tag{9}$$

Note the denominator ${}_k \ |\mathcal{W}_i^k|^2 = |k| * h^2$ is a constant. Intuitively, the similarity between diseases $i$ and $j$ is higher when two diseases visit more shared nodes at a higher frequency.

### S3 Note 3: Further details about TxGNN explainer

A machine learning model can provide accurate disease treatment predictions. However, for domain scientists' adoption, prediction alone is not sufficient. Thus, a model is expected to generate why it outputs this prediction in a form familiar to domain experts' decision-making. In the case of treatment prediction, an ideal form of explanation is to simulate how drug developers approach drug-disease relation — that is, to understand how a drug perturbs the local biological system such that it creates a therapeutic effect on the disease pathway. As TxGNN leverages the large-scale biological knowledge graph, we can probe into the local neighborhood around a query drug-disease node and pinpoint the exact mechanism contributing to the prediction. However, as a biological network is complex, making meaningful explanations requires a model to prune most uninformative edges and extract a sparse version of the local neighborhood. This can be formulated as a graph explainability problem where we try to identify a sparse set of edges where the model can make a faithful prediction using these edges[27]. To achieve it, we develop a post-training graph explainability module, adapted from GraphMask approach[28], that can drop spurious edges from the dataset and retain a sparse set of edges that contribute most towards the prediction.

**Necessary adaptations of GraphMask approach for biomedical knowledge graphs.** We modify GraphMask[28] in the following manner to generate meaningful local explanation subgraphs of the knowledge graph. (1) Instead of a complex gate that outputs scores close to 0/1, we adopt a smooth sigmoid gate where predictions are uniform across 0 to 1. This is important because we find hard concrete map edges to 1 as long as they affect the model prediction. However, this still keeps many edges that preclude us from making acceptable medical explanations. Instead, the sigmoid gate allows us to distinguish the intensity of contributions and provides a flexible framework. By setting a threshold, we remove large amounts of positive edges and only retain ones crucial for the model prediction. (2) Second, while GraphMask has a single learnable weight for every edge in the dataset, we adopt a separate weight for each relation. Since embeddings across relations are different, the model assigns uniformly high scores for all edges of a given relation type despite edges varying in relevance. Using relation-specific weights allows the model to capture the importance scores of individual edges.

### S4 Notes 4: Experimental setup and implementation details

Next, we outline the experimental setup, including information on performance evaluation and dataset splits. We also provide details on the practical implementation of TxGNN deep graph models.

#### S4.1 Creating dataset splits for rigorous performance evaluation

Our dataset presents well-studied information and includes the vast majority of existing treatments. As a result, it is easy to predict treatments for diseases with various pre-existing treatments. However, for zero-shot prediction of therapeutic use, we need to make good predictions on conditions with few or no current treatments available. The classical random split of edges of the knowledge graph into training and testing sets would not simulate this application. In the random split, for diseases with many known indications, the model would view some of these drug-disease edges in training and thus easily predict therapeutic use based on drug similarity. However, this would prevent the model from assimilating meaningful biological knowledge. Therefore, we consider the following dataset splits into training and test sets:

- **Disease area splits:** Many diseases of therapeutic interest have no existing treatments and lack significant biological knowledge. To evaluate whether TxGNN would be robust to predicting drug-disease relationships for such diseases, we develop data splits that simulate well-studied diseases as molecularly uncharacterized diseases. We cannot directly test on molecularly uncharacterized diseases, such as rare diseases, because the treatments are too few to build a confident machine learning model. We select nine disease groups: cell proliferation, mental health, cardiovascular diseases, anemia, adrenal gland, diabetes, autoimmune, neurodegenerative, and metabolic disorders, and then extract groups for these diseases from the Disease Ontology hierarchy such that the group includes the disease and all its children. Since these well-studied diseases have many drug-disease relationships, we can easily evaluate the model's performance during the simulation.

  For each disease, we create a separate data split as follows. First, all the drug-disease edges connected to the diseases in the group are moved to the test set. As a result, TxGNN has no information about existing indications and contraindications use edges for the chosen disease group during training. This simulates the lack of existing treatments encountered with molecularly uncharacterized diseases. Next, we remove a significant fraction of the local 1-hop subgraph neighborhood for the disease group. Again, this simulates the limited biological understanding of molecularly uncharacterized diseases.

- **Systematic dataset splits:** The deployed machine learning model should excel at predicting diseases without known treatments. Predicting new treatments for diseases that already have treatments is easier than predicting diseases without treatments. This is because information about existing treatments can directly illuminate the molecular mechanism, and drug similarity can help infer new treatments. Thus, to robustly test our model, we design this split to systematically study prediction on novel unseen diseases. To do that, we first randomly split the entire set of diseases. Then, we take all drug-disease relations associated with the testing set of diseases to the test set such that there are no known treatments during training and the testing set consists of novel diseases. The testing set has around 100 different diseases in each randomly seeded run.

##### S4.2 Modeling molecular and clinical relationships

In graphs, each edge typically has a direction and points from the source to the target node. However, in our biological knowledge graph, edges are bidirectional. For example, a drug $A$ indicated for disease $B$ is represented in TxGNN by a tuple $A$, indication, $B$. Similarly, disease $B$ can be treated by drug $A$, corresponding to a tuple $B$, rev_indication, $A$. For homogeneous relation type (*e.g.*, protein-protein interactions) where the head and tail node belong to the same node types, there is no additional reverse relation type as the reverse edges are collapsed into the original relation type. Thus, we add these reverse relation types to the knowledge graph, following standard practice[29]. For the sake of notation, when the reverse relation has a different relation type from the original type $r$, we denote the reverse relation type as $r^c$.

##### S4.3 Negative sampling for training TxGNN models

As we only have positive data, negative data are constructed via sampling. The sampling from the unobserved simulates the realistic constraint where most possible drugs do not interact with the disease. For each relation type, we fix the source nodes and permute the target nodes through either random sampling from the set of nodes associated with this relation type's target nodes or a weighted sampling based on the degree of the target nodes. As we conduct reverse relation type construction, the source node type would also be shuffled and included in the negative samples when we do sampling for the reverse relation type. This concept of negative sampling based on shuffling target nodes is crucial. For example, suppose we want to study drugs $A$ that can treat disease $B$, then we narrow down to the relation $B$, rev_indication, $A$ instead of the $A$, indication, $B$. At each training iteration, we conduct a different set of negative samples of equal size to the number of positive samples.

##### S4.4 Hyperparameter tuning

We conduct hyperparameter tuning using Hyperband on validation set micro AUROC using complex disease split following two stages. The first is to optimize the parameters for pre-training and fix fine-tuning parameters, where we conduct a sweep of grid search with a learning rate of $\{1e-4, 5e-4, 1e-3\}$, batch size of $\{1024, 2048\}$, and epoch size of $\{1, 2, 3\}$. Next, we fix the pre-training parameters and do a grid search for fine-tuning parameters with the hidden size of $\{64, 128, 256, 512\}$, input size of $\{64, 128, 256, 512\}$, output size of $\{64, 128, 256, 512\}$, number of inter-disease prototypes of $\{3, 5, 10, 20, 50\}$ and learning rate of $\{1e-4, 5e-4, 1e-3\}$. We obtain a final set of hyperparameters with a pre-training learning rate of $1e-3$, batch size of $1024$, epoch size of $2$, fine-tuning learning rate of $5e-4$, hidden size of $512$, input size of $512$, output size of $512$, number of prototypes $3$.

##### S4.5 Implementation details

The TxGNN is implemented using DGL[30] and PyTorch[31] Python deep learning frameworks. We use Pandas[32], Numpy[33] for data processing and computing; scikit-learn[34] for evaluation metrics; seaborn[35], matplotlib[36], UMAP[37] for visualization; Weights and Bias (https://www.wandb.ai) for training monitoring and hyperparameter tuning. We train the model with one NVIDIA Tesla V100 GPU in a server. TxGNN Explainer is implemented in JavaScript using React.js[38], D3.js[39], and Ant Design[40]. The graph data is managed using Neo4j database[41]. TxGNN Explainer communicates with TxGNN through a Python web server built with Flask[42].

##### S4.6 Further information on existing methods and implementation details

We note that KL/JS/DSD/Proximity are network science heuristics calculated from the network properties based on proximity principles. They are not learnable and, thus, are not optimized for the prediction tasks and underperforming learnable methods. RGCN uses the same backbone as TxGNN without our proposed metric learning and pretraining modules. From this comparison, we see the utility of the metric learning module. HGT and HAN are state-of-the-art KG message-passing methods shown in graph machine-learning literature to be more expressive than RGCN. However, as our experiments show, they typically perform similarly to RGCN. Also, they are expensive to run compared to RGCN. Thus, we chose RGCN as the model backbone. BioBERT is a pure text-based approach that feeds disease and drug descriptions into a large biomedical language model. Thus, they utilize orthogonal information compared to the network approach. We see that BioBERT is better than most network-based methods. This is because of our data split construction where we explicitly remove links for the testing diseases, but this removal does not affect the textual information in the BioBERT model. But it still underperforms compared to TxGNN, highlighting the effective predictive power. In addition, we observe that BioBERT has significant performance drops in several disease area splits since some disease areas may lack literature information, which is a disadvantage of the text-based approach. Integrating a text-based approach with network methods is feasible, but we leave it as future work.

Regarding implementations, we follow the original author's implementations for the baselines. Particularly, for network medicine statistics including KL, JS, proximity, and DSD, we used the codebase that recently benchmarked these scores for COVID-19 targets in https://github.com/Barabasi-Lab/COVID-19/tree/main. We used HANConv and HGTConv layer implementation in the Pytorch Geometric library for HAN and HGT. For BioBERT, we used the huggingface repository https://huggingface.co/dmis-lab/biobert-v1.1 to download the raw model weights and then applied an MLP decoder to make predictions. For all baselines, we use the exact same data splits as in TxGNN for a fair comparison.

#### Supplementary Figures

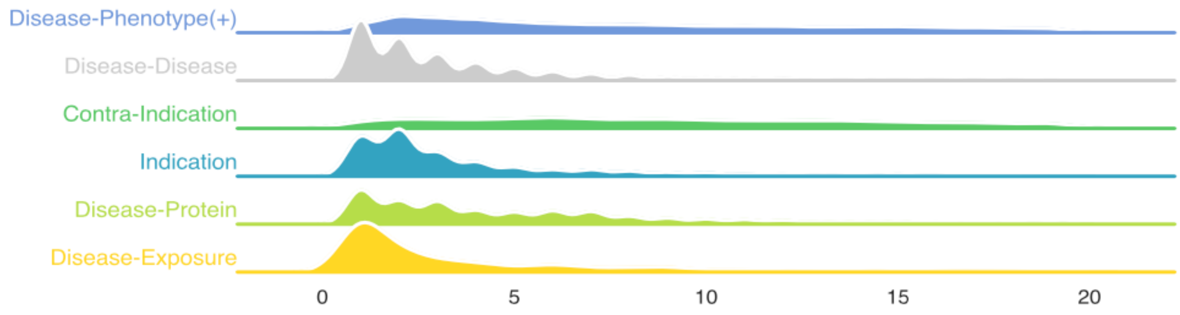


**Figure S1: Distribution of the number of relationships incident to diseases in the medical knowledge graph (KG).** Our KG has rich information about diseases. On median, for diseases that have known indications, a disease node is connected to 5 proteins, 14 phenotypes, 3 other diseases, and 2 exposures in the network.

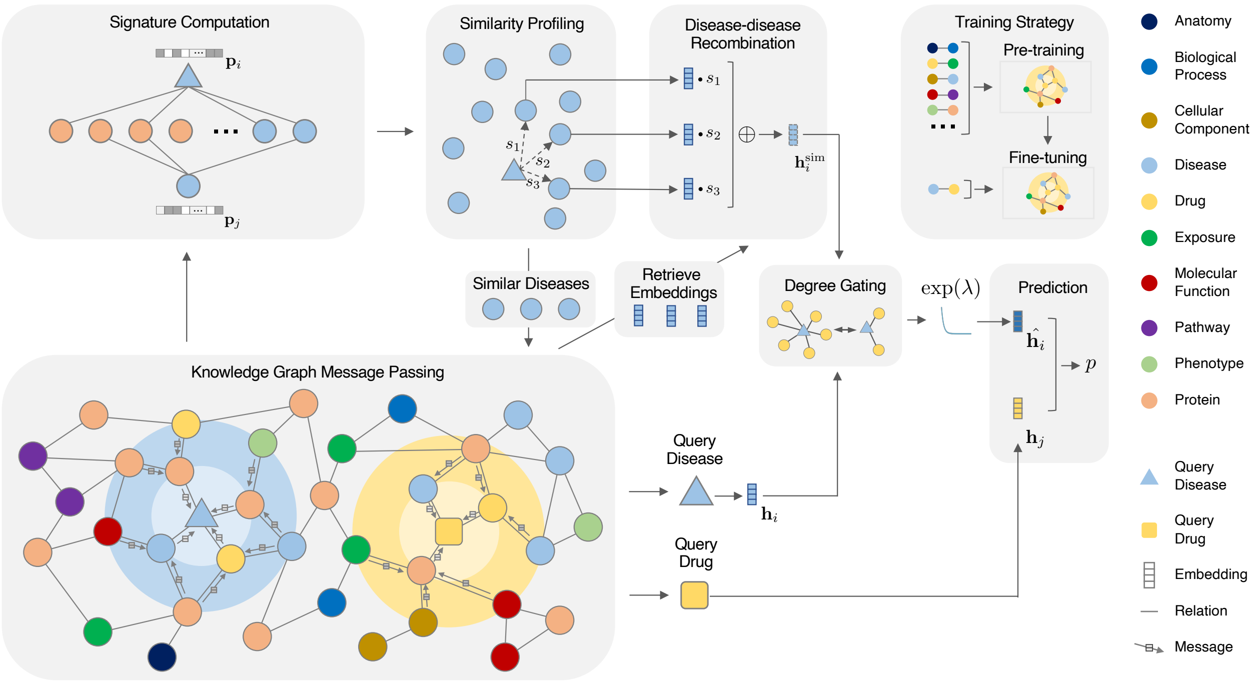


**Figure S2: Overview of TxGNN Predictor**. (1) TxGNN projects biological concepts into meaningful representations through knowledge graph neural network message passing on the KG. (2) It then designs a similarity disease search component to enrich molecularly uncharacterized diseases and has three modules (2.1) It computes a signature vector for each disease that captures the disease similarity. (2.2) Based on the signature vector distance, it profiles a set of similar diseases and retrieves their latent embeddings. (2.3) It then aggregates similar diseases into a powerful auxiliary embedding. (2.4) A gating mechanism is designed to control the effect between the original and auxiliary disease embedding since many well-characterized diseases have sufficient embeddings and do not need subsidies. (3) A decoder then maps the query drug and disease representation to predict the outcome. A pretext learning stage is devised to allow TxGNN to learn an initialized embedding that captures complex biological knowledge.

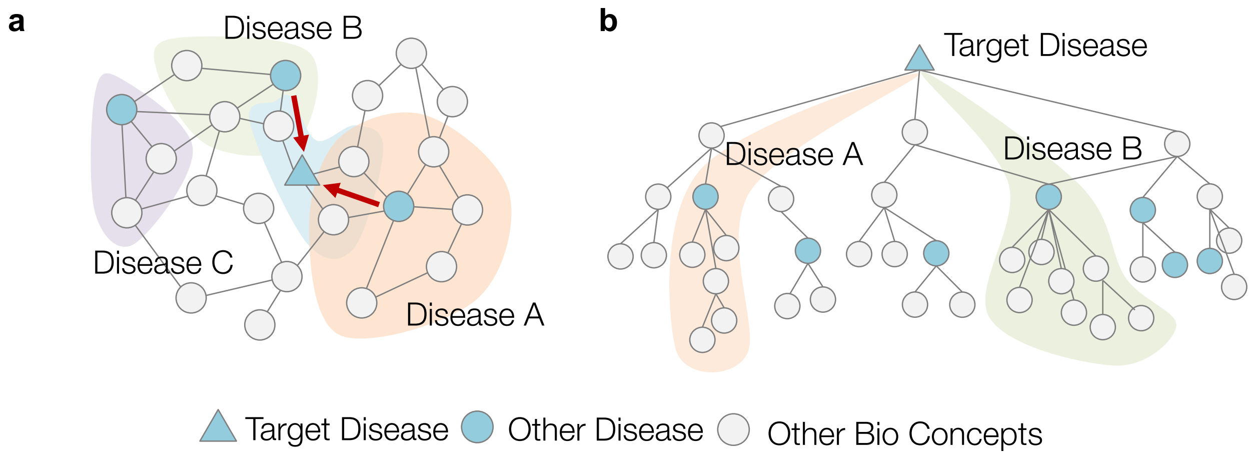


**Figure S3: Illustration of TxGNN graph rewiring perspective. a.** For a target disease, we use a disease signature to select similar disease nodes in the entire network (in the figure, the target disease selects diseases A and B, not C). We then want to aggregate the disease module information of these similar disease nodes into the target disease, where this information would not be available with the classic GNN approach. TxGNN fuses this information into target disease embedding by aggregating these remote disease embeddings to the target disease embeddings. After fusing, the disease module information is available in the target disease node. **b.** This fusing step happens in the latent embedding space, which is equivalent to adding a network edge between the target disease node and selected similar diseases. We see that through this network rewiring perspective, TxGNN conducts a long-range selective aggregation guided by domain prior.

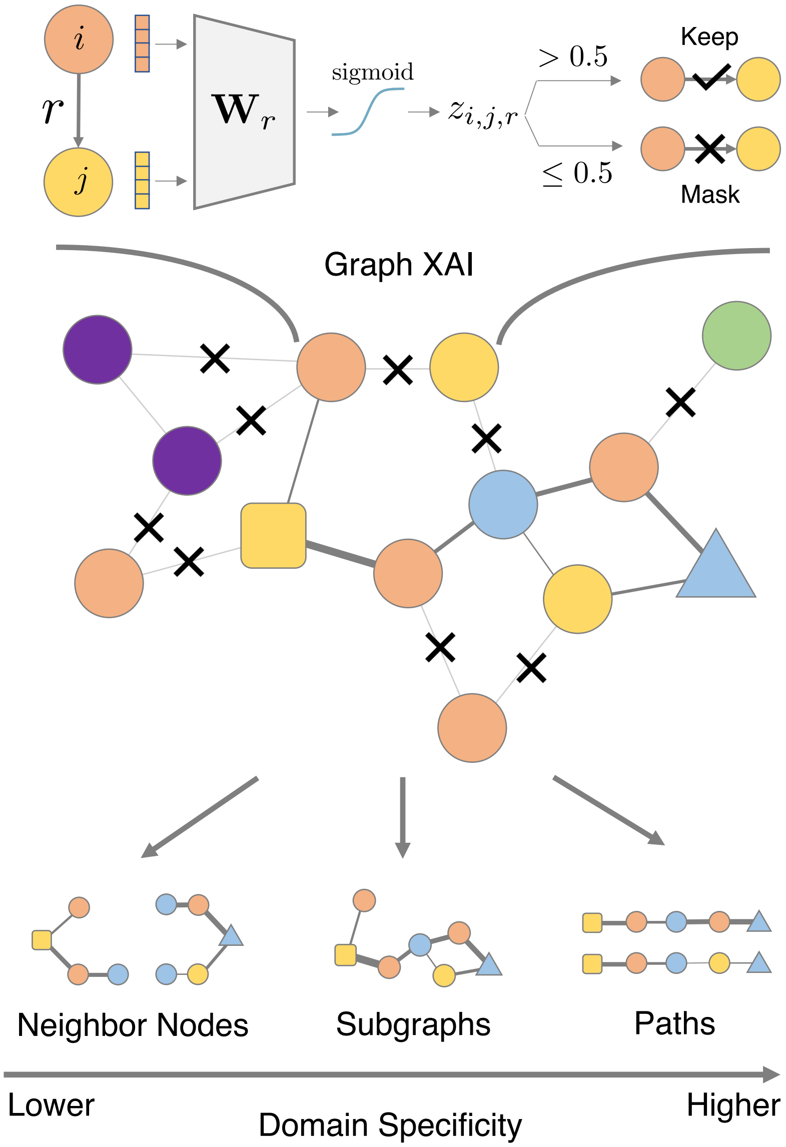


**Figure S4: Overview of TxGNN Explainer.** For each edge between node $i$ and $j$ with relation $r$, a weight matrix $\mathbf{W}_r$ takes in the node embedding and produces a score $z_{i,j,r}$. If the score measures the importance of this edge to the prediction. If it is larger than the user-defined threshold, the edge is kept and deleted otherwise. Applying this to every edge, we can obtain a sparse subgraph that depicts the essential connections for predictions. This same subgraph explanation is then converted to various forms of visualizations such as neighbor nodes, subgraphs, and paths. We have studied path-based visualization and found it aligns best with human experts[43].

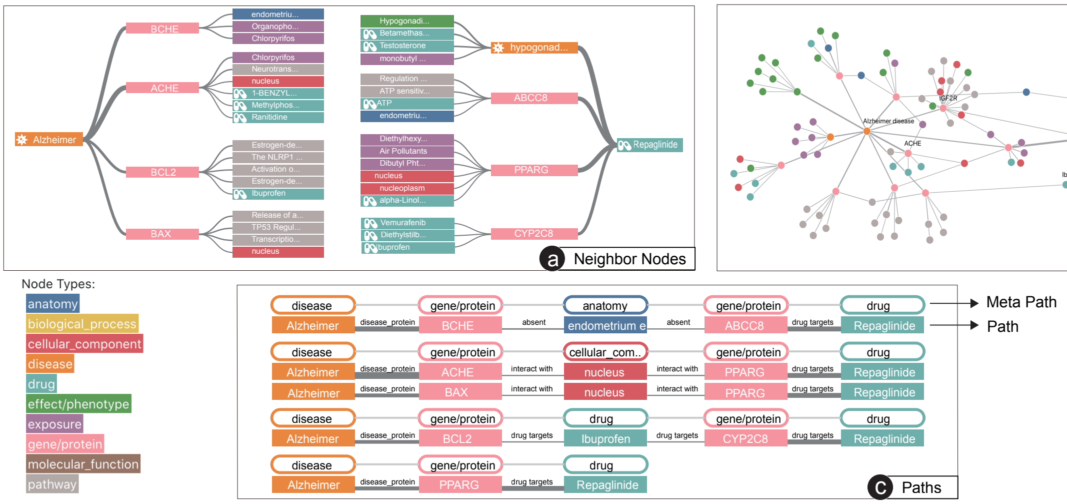


**Figure S5: Three types of visual explanations for TxGNN predictions.** We categorize instance explanations for GNNs into three main groups, neighbor nodes (a), subgraphs (b), and paths (c). We compared the three visual explanations and selected path-based explanations due to their similarity to the clinicians' reasoning about indications.

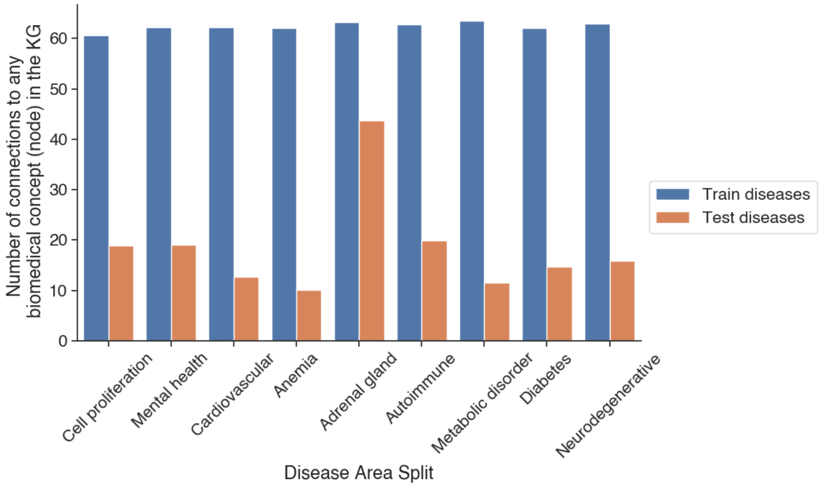


**Figure S6: Number of first-hop neighbors for testing and training set diseases in the disease area splits.** To show that TxGNN works for diseases with limited knowledge connections to KG, we visualize the number of connections (first-hop neighbors) for testing and training set diseases in the disease area splits. In the figure above, we see a significantly smaller number of connections to the underlying KG for testing diseases, thus suggesting that we are already evaluating diseases with limited knowledge compared to other average diseases. For example, for cell proliferation split, testing diseases, on average, have 18.8 connections to the KG whereas training diseases have 60.6 connections, accounting for 3.22 times smaller connections. For example, in the Anemia disease area split, testing diseases, on average, have 10.1 connections to KG and training diseases 62.1, with a 6.17 times smaller number of connections.

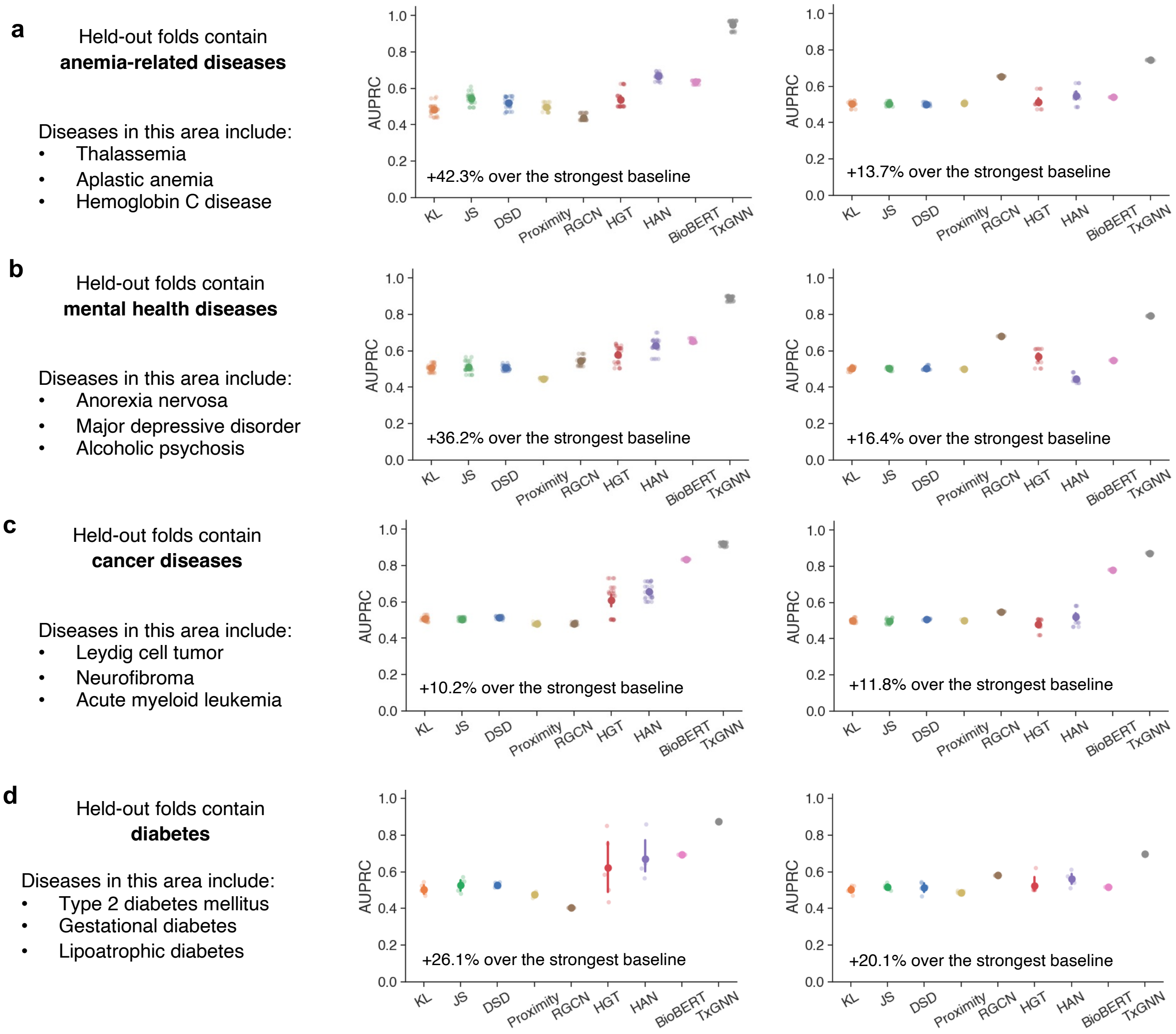


**Figure S7: Additional benchmarking results of TxGNN. a-d.** We included four disease areas: anemia, mental health, cancer, and diabetes. We observe consistently robust performance of TxGNN compared to baselines. The evaluation utilizes the area under the precision-recall curve (AUPRC) and is conducted with five random data splits (N=5). The mean performance is highlighted, while the 95% confidence intervals are represented by error bars.

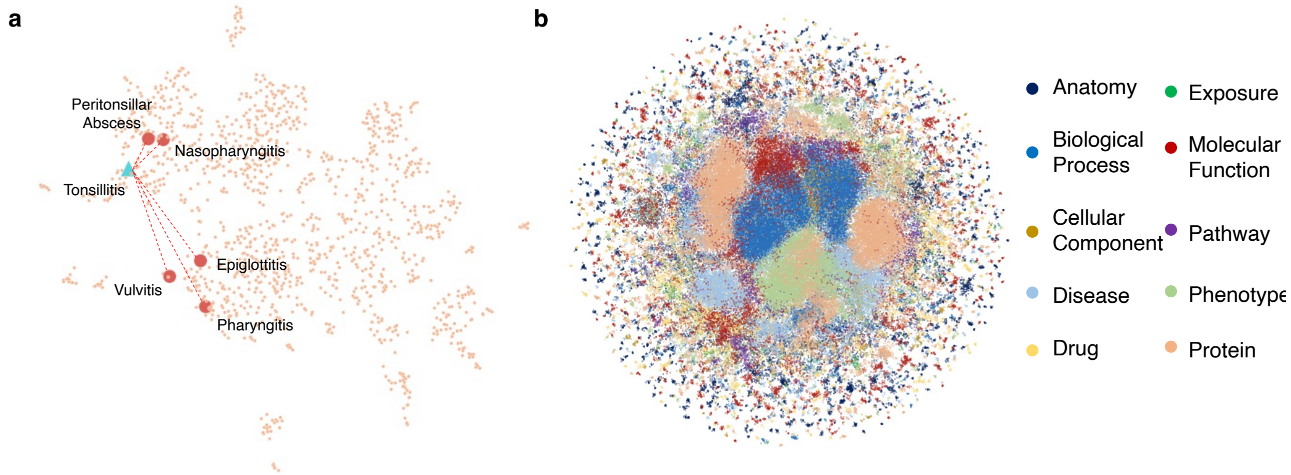


**Figure S8: Visualizing TxGNN's latent representations. a.** To further understand the performance gain of the metric learning module from a machine learning standpoint, we explore the example of tonsillitis. Diseases similar to tonsillitis (epiglottitis, peritonsillar abscess, nasopharyngitis, pharyngitis, vulvitis) are initially distant in the embedding space. Thus, by fusing distant disease embeddings, TxGNN establishes a long-range skip connection to the disease module of these similar diseases and provides complementary information missing from the local neighborhood around the target disease. This is especially beneficial in predicting therapeutic use for conditions with few or no treatments and limited molecular understanding. TxGNN uses disease signatures as a learnable disease look-up catalog to identify the appropriate distant disease information that can be transferred to the underpowered target disease. **b.** Visualization of embeddings for all nodes in the KG (Silouette score: 0.2647, Adjusted Rank Index: 0.2188).

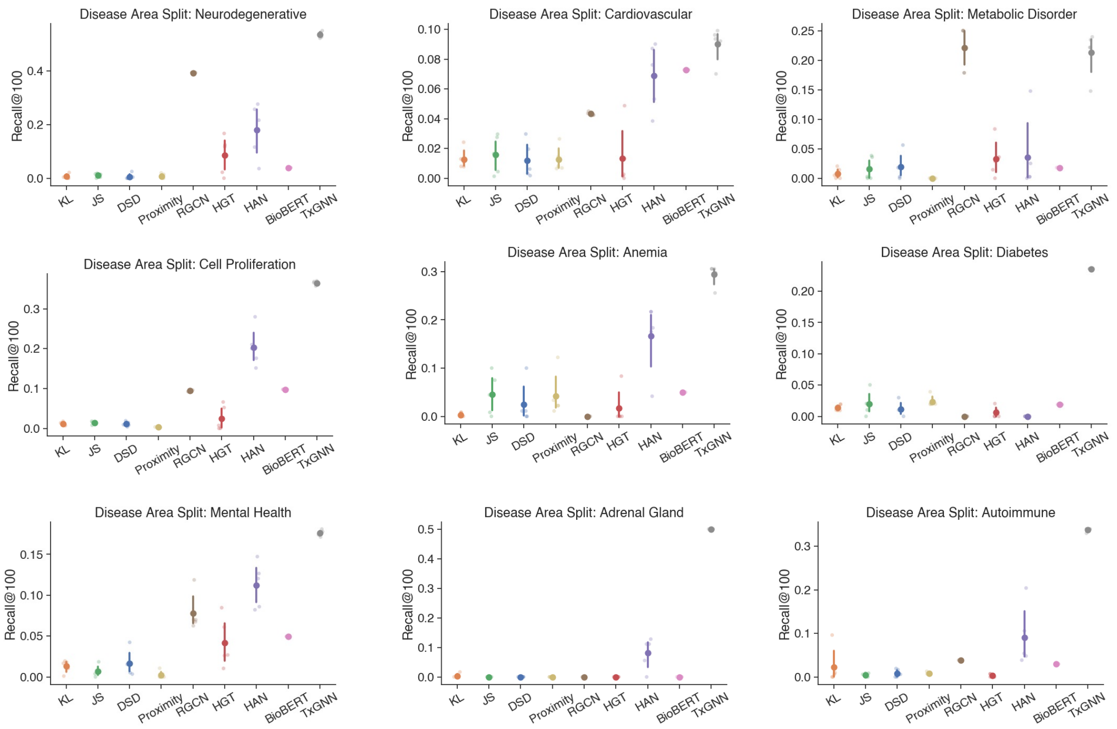


**Figure S9: Benchmarking indication predictions using Recall@100 performance metric.** We generate a list of the top-100 predicted drugs for each disease and calculate the fraction of correct hits in the list (Recall@100). TxGNN performs consistently better across hold-out sets. This evaluation is conducted with five random data splits (N=5). The mean performance is highlighted, while the 95% confidence intervals are represented by error bars.

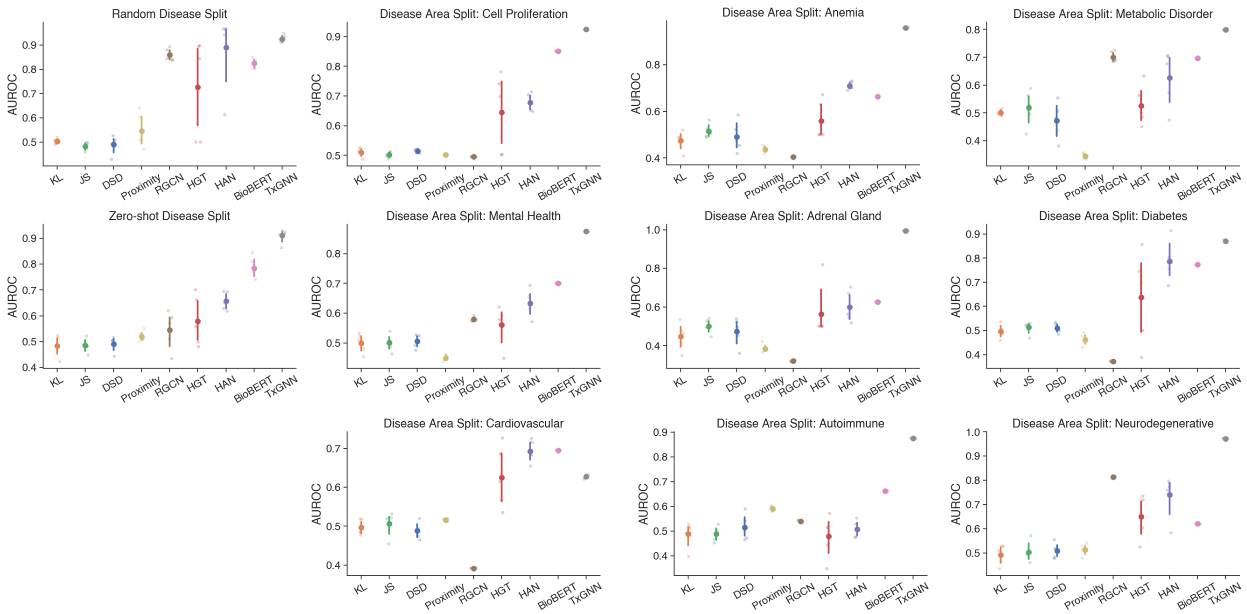


**Figure S10: Benchmarking indication predictions using AUROC performance metric.** AUROC metric for all disease splits under indication prediction in TxGNN. We observe consistent performance improvement. This evaluation is conducted with five random data splits (N=5). The mean performance is highlighted, while the 95% confidence intervals are represented by error bars.

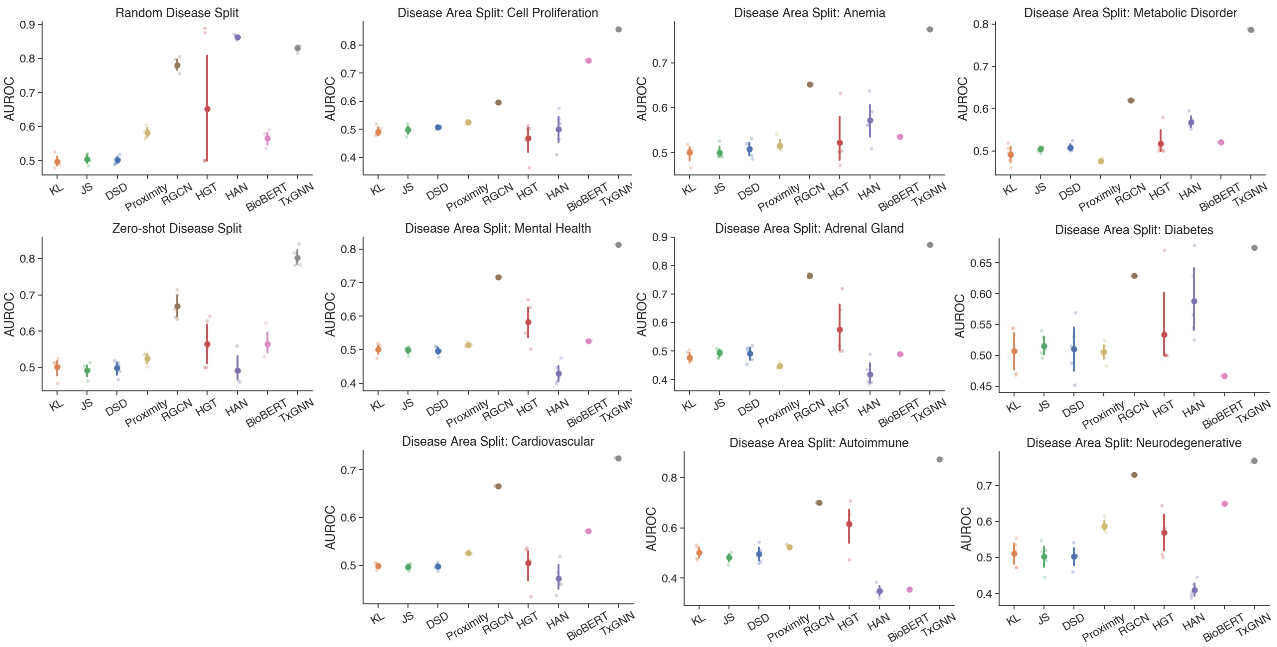


**Figure S11: Benchmarking contraindication predictions using AUROC performance metric.** AUROC metric for all disease splits under contraindication prediction in TxGNN. Results show consistently strong performance of TxGNN and improvement over baseline methods. This evaluation is conducted with five random data splits (N=5). The mean performance is highlighted, while the 95% confidence intervals are represented by error bars.

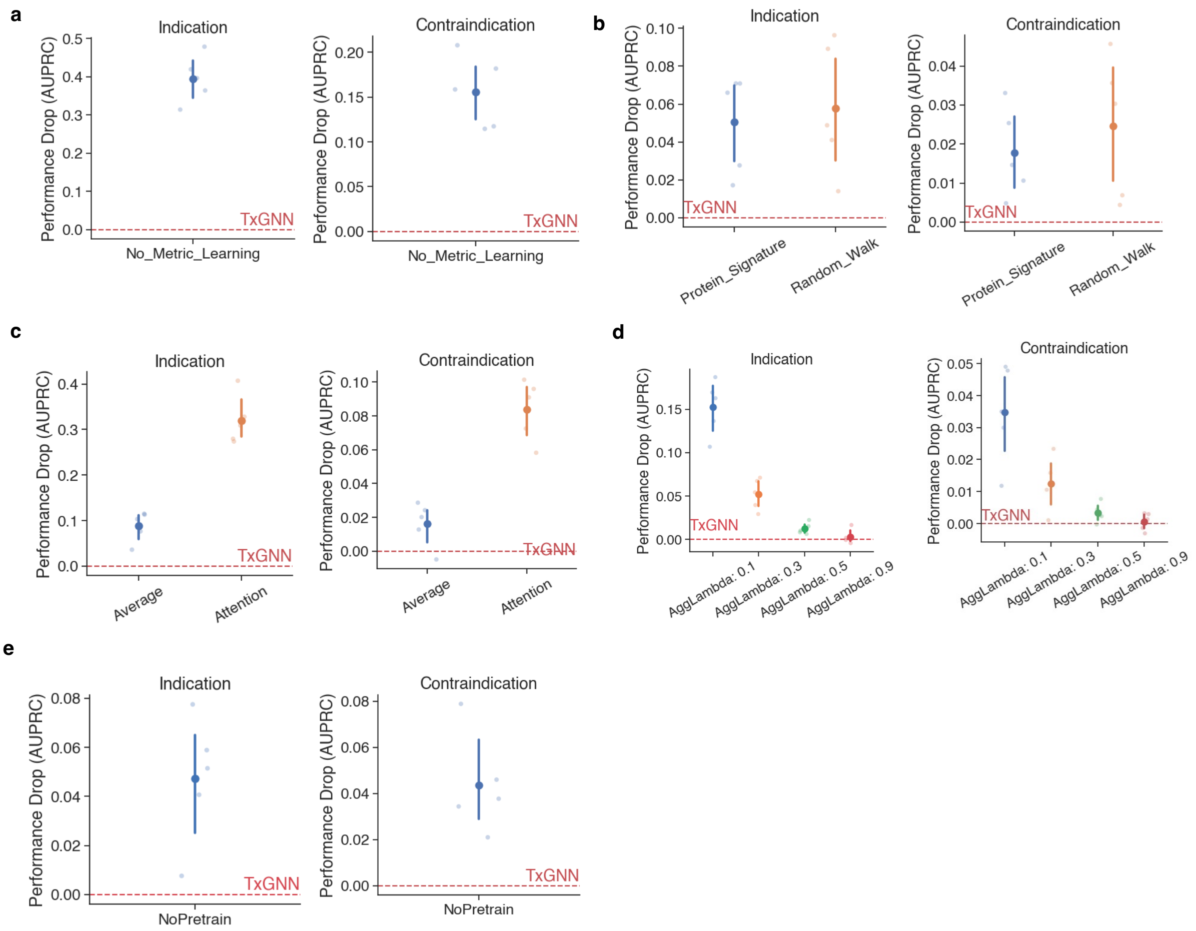


**Figure S12: Ablation analyses.** We have conducted five ablation studies. The key module is the disease-disease metric learning module where we update the target disease embedding through similar diseases. **a.** In the first ablation, we remove this module entirely, using the raw KG embedding from the query disease to make the prediction. As we see from the results, the performance drops to 0.4 AUPRC, suggesting the overall utility of the metric learning module. Next, we delve into individual components of the metric learning module. Notably, three components have alternative options. **b.** The first one is the choice of disease similarity signature. We experimented with protein-based signatures and also random walk-based signatures. We observed performance drops compared to the TxGNN, showing the great performance of the current signature where we utilize all node types linked to the disease. **c.** Another component is the degree-based gating module, which dynamically adjusts the embeddings from the metric learning module or the original disease embedding module. The intuition is to have a higher weight in the metric learning module when there is less information about the diseases and a lower weight and vice versa. We use an exponential function to achieve this. We first test if this degree-based weighting is necessary by introducing other weighting mechanisms like attention scores or simply taking an average. We observe that the degree-based mechanism has significantly better performance. **d.** We then test the performance impact when we adjust this exponential parameter in the degree-based weighting mechanism. TxGNN (AggLambda = 0.7) performs significantly better when compared with 0.3 and 0.1 and is on par/slightly better with 0.9. e. Last but not least, we also introduce a pre-training module to learn from the broad biology contained in the KG. We thus remove the pre-training module. All evaluations from **a**-**e** are conducted with five random data splits (N=5). The mean performance is highlighted, while the 95% confidence intervals are represented by error bars.

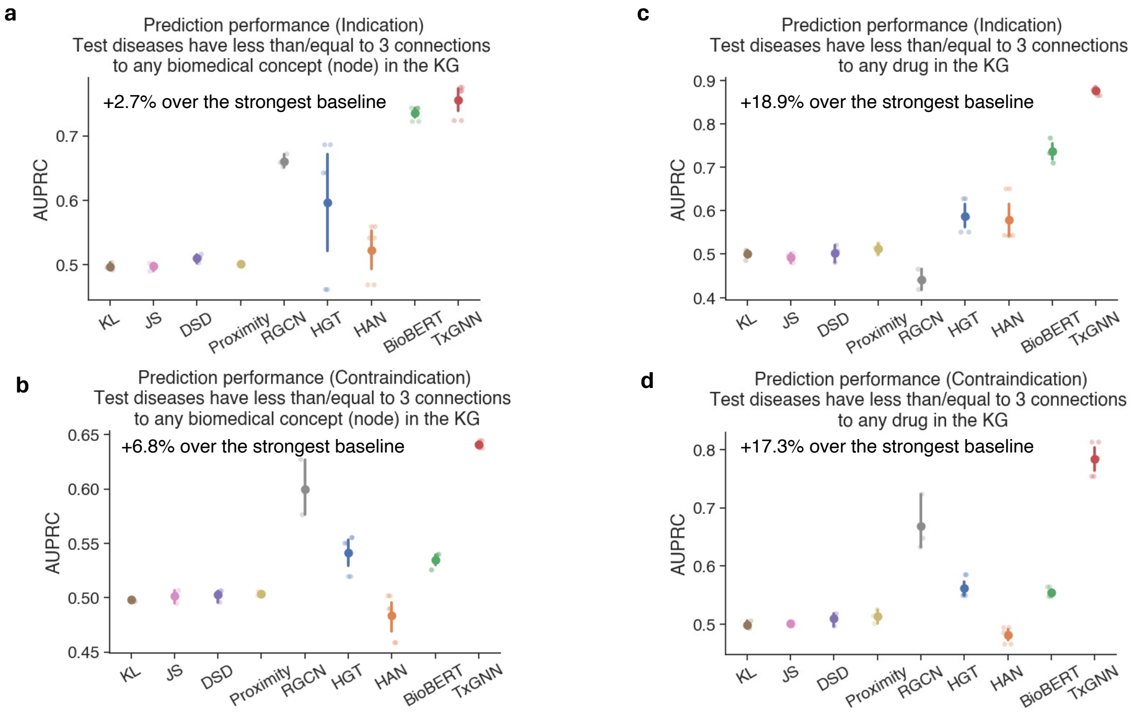


**Figure S13: Additional evaluation of TxGNN in settings with extremely scarce disease data.** While existing data splits showcase TxGNN prevails over these realistic settings, we construct two new data splits to further test TxGNN in challenging settings when diseases have an extremely small number of connections to KG. **a**-**b.** Firstly, we move diseases with fewer than 3 disease relationships in the KG to the test set. Note that this is an extremely difficult setting that corresponds to diseases with minimal molecular understanding, and the model is asked to make predictions by relying only on fewer than 3 disease relationships. **c**-**d.** Secondly, we move diseases with <=3 number of indications to the test set and report their performances. In both settings, we observe consistent improvement of TxGNN over all existing models. All evaluations from **a**-**d** are conducted with five random data splits (N=5). The mean performance is highlighted, while the 95% confidence intervals are represented by error bars.

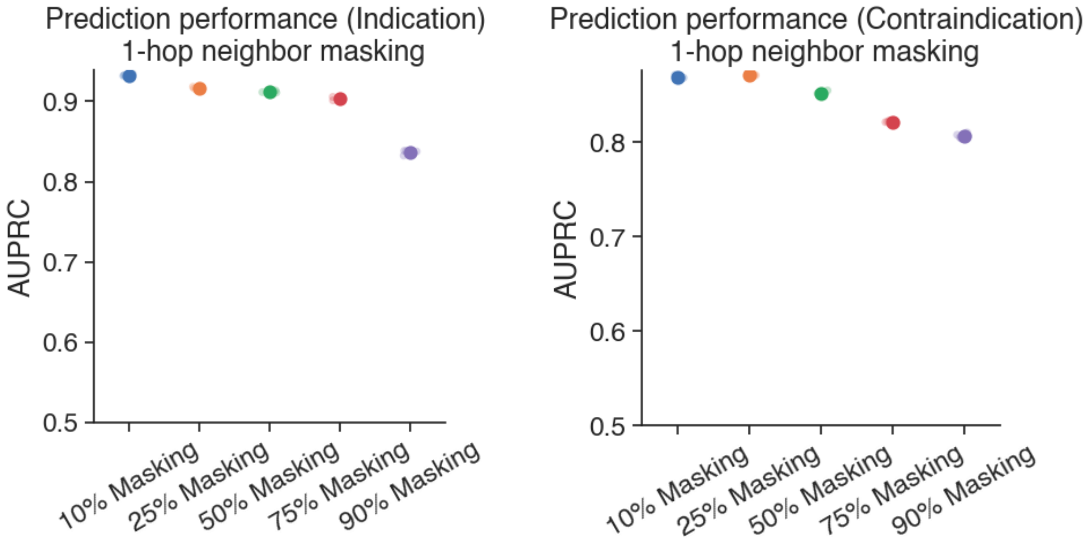


**Figure S14: Robustness of TxGNN to scarce disease information.** We study how the TxGNN's performance changes when varying amounts of direct disease information are removed from the model. To show it, we conducted an experiment where we removed 10%, 25%, 50%, 75%, and 90% of one-hop neighbors around these testing diseases in a disease area for TxGNN. We observe stable performance despite an expectedly decreasing trend (for indication, 1.64%/2.19%/3.02% in 25%/50%/75% of mask ratios, and for contraindication, 1.14%/4.20%/7.2% in 25%/50%/75% of mask ratios), suggesting TxGNN 's robustness. When we mask the 90% of 1-hop neighbor, the performance degrades from 0.932 AUPRC to 0.837 AUPRC for indication and from 0.863 AUPRC to 0.791 AUPRC for contraindication, suggesting a still accurate predictive model even under severe information removal.

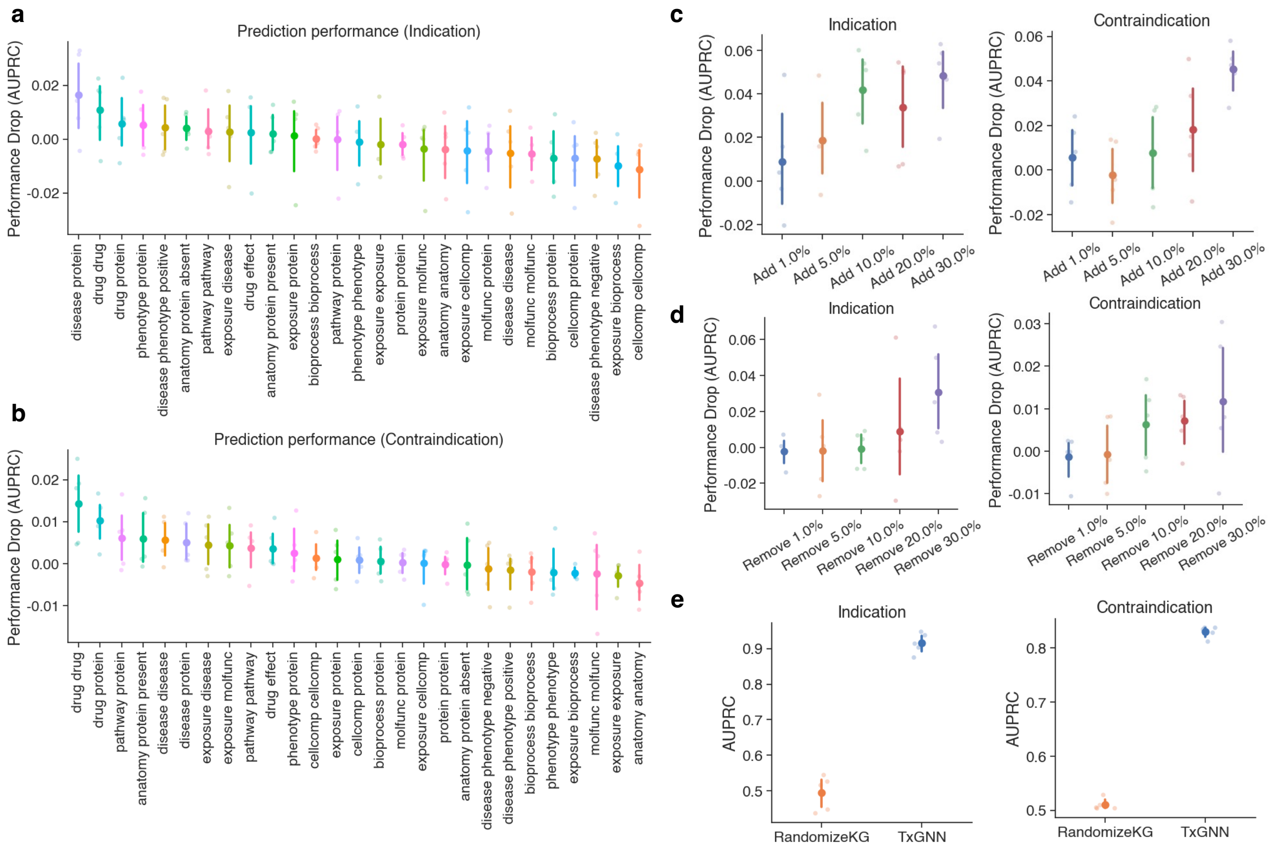


**Figure S15: Robustness of TxGNN to incomplete and noisy medical knowledge graph (KG).** We conducted three alteration types on the KG to evaluate the robustness of TxGNN. **a-b.** Firstly, we remove all edges in an edge type in the KG to test the contribution of this edge type to the model. **a.** We find that overall TxGNN performance is stable against edge-type removals. The maximum performance drop is 0.018 AUPRC for indication and 0.015 AUPRC for contraindication. **b.** The most significant performance drop corresponds to the most important edge types. Here we highlight a few important observations. For indication, the top edge type is disease protein, and the third important edge type is drug-protein, which makes intuitive sense since the drug acts on target proteins in the disease for indication. Phenotype-protein and disease-phenotypes are important for indications since phenotype corresponds to shared disease molecular mechanisms. Drug-drug interactions are an important edge type for both contraindications (rank 1) and indications (rank 2). This is due to similar drugs having similar indications and contraindications. **c-d.** The second ablation is to randomly perturb part of the KG and see performance changes. We randomly add 1%/5%/10%/20%/30% of edges to the KG and randomly remove 1%/5%/10%/20%/30% of edges from the KG and report performance differences. We report a few interesting observations. First, TxGNN is robust to random perturbations in the KG. Second, as expected, the performance decreases as the removal/addition rate gets higher. Interestingly, removing up to 10% of edges does not affect TxGNN's performance much, suggesting that numerous edges are not used in indication predictions, corroborating the usefulness of the XAI method in prioritizing important paths. **e.** Last, we randomize the KG by permuting each edge index within an edge relation. We find that by randomizing KG, the performance decreases significantly, showing that the KG contains crucial information that enables model predictions. All evaluations from **a-e** are conducted with five random data splits (N=5). The mean performance is highlighted, while the 95% confidence intervals are represented by error bars.

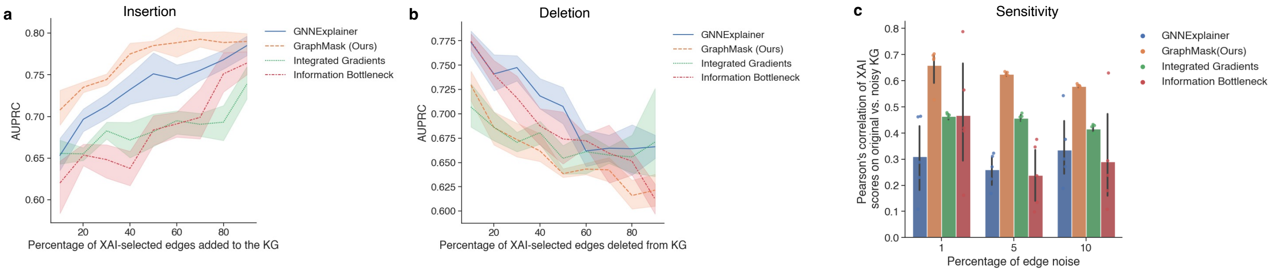


**Figure S16: Benchmarking explainability (XAI) algorithms and TxGNN Explainer.** We benchmarked graph XAI algorithms using the following three metrics: **a. Insertion (faithfulness)**: we start from an EMPTY graph and gradually INSERT top K% edges to the graph where the importance is measured by each graph XAI method. We measure the updated AUPRC on the test set using different K. A better XAI method has a higher AUPRC at each threshold since it suggests that the newly added edges are informative to the graph predictions. Note that this definition is also commonly referred to as fidelity+ in the graph XAI literature. **b. Deletion (faithfulness)**: we start from the FULL graph and gradually DELETE top K% edges to the graph where the importance is measured by each graph XAI method. We measure the updated AUPRC on the test set using different K. A better XAI method has a lower AUPRC at each threshold since it suggests that the deleted edges are necessary for the graph predictions. Note that this definition is also commonly referred to as fidelity- in the graph XAI literature. **c. Sensitivity (stability)**: we randomly add X% of edges to the graph and rerun the graph XAI algorithm to produce explanations. We then compute the Pearson correlation between the base and perturbed explanations. A larger Pearson correlation suggests stability of explanation against perturbations. Overall, we observe that our method (GraphMask) has better results. Notably, top-ranked explainable edges in GraphMask are shown to have the most essential edges when removed from a full graph or inserted in an empty graph. The performance is also robust across all insertion/deletion percentages. These two results show the faithfulness of GraphMask and that it can retrieve relevant edges to explain model predictions. In addition, GraphMask is relatively stable when under perturbation of the KG. All evaluations from **a-c** are conducted with five random data splits (N=5). The mean performance is highlighted, while the 95% confidence intervals are represented by error bars.

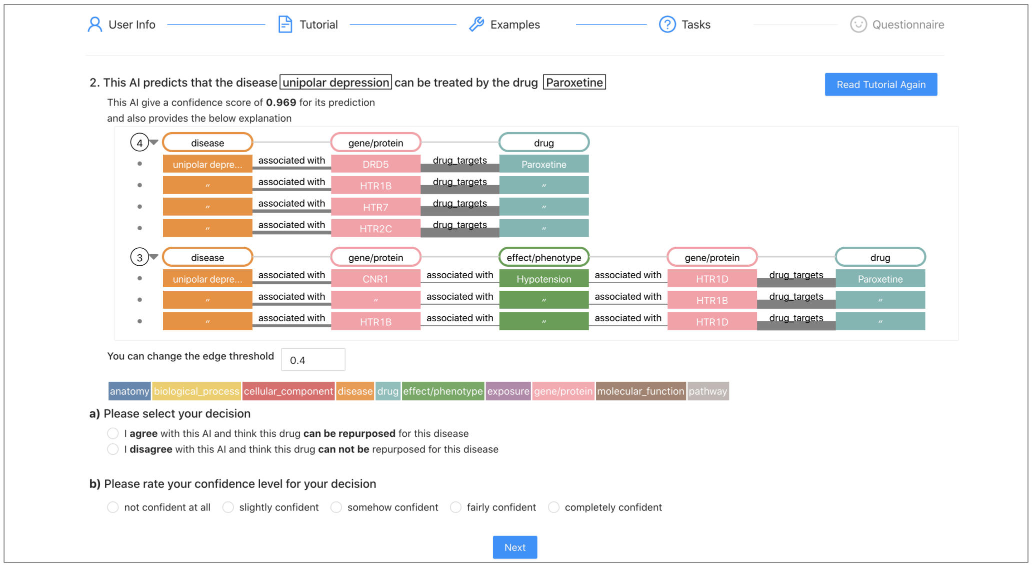


**Figure S17: Interface used in the usability study of TxGNN.** We compare path-based explanations with a non-explanation baseline. For each prediction, participants decided whether the predicted drug could treat a certain disease and reported their confidence levels using a 5-point Likert scale (1=not confident at all, 5=completely confident).

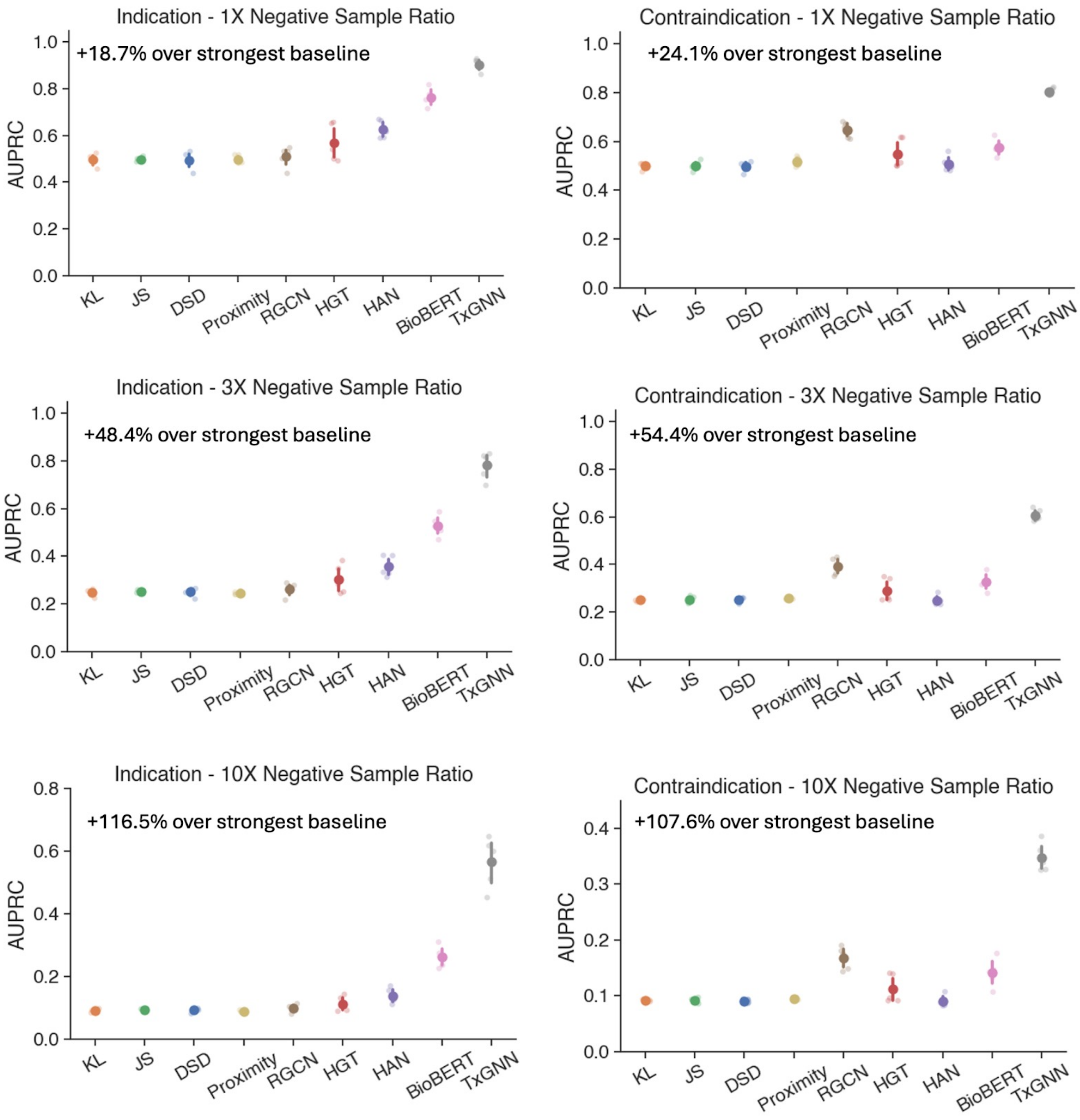


**Figure S18: Additional benchmarking of TxGNN under varying numbers of negative samples.** We vary the negative samples in the testing set (1/3/10 times the positive samples). We observe a consistent improvement of TxGNN over baselines under various negative sample ratios. Higher negative sample ratios also lead to a more pronounced improvement of TxGNN. All evaluations are conducted with five random data splits (N=5). The mean performance is highlighted, while the 95% confidence intervals are represented by error bars.

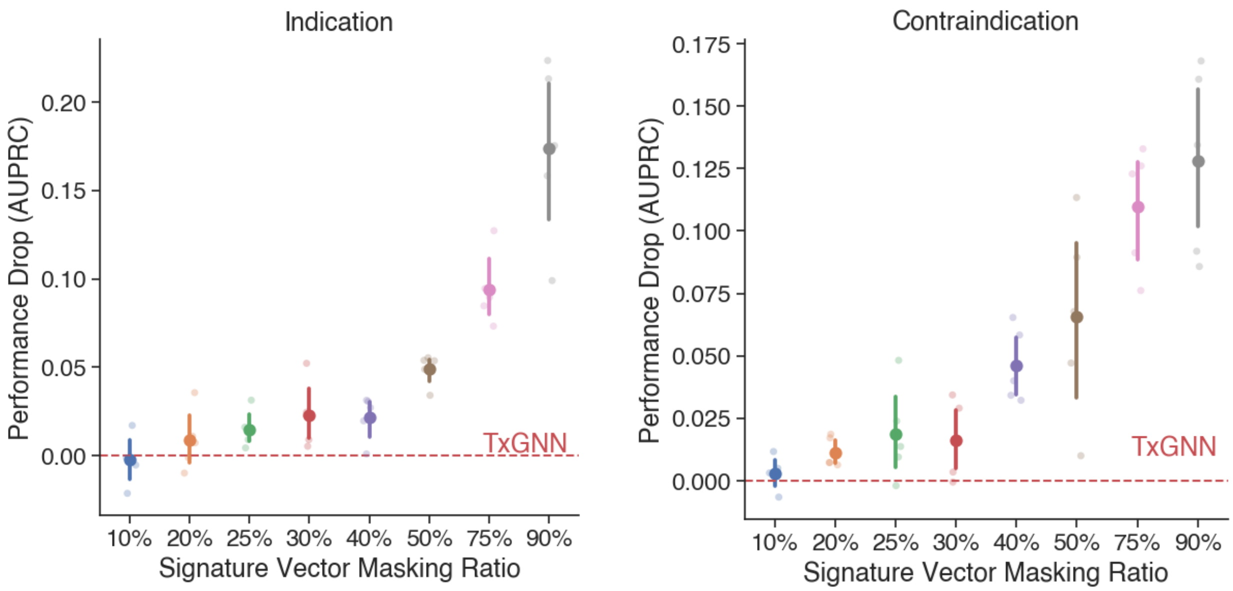


**Figure S19: Robustness of disease signature vectors in TxGNN.** We remove fractions of disease signature vectors in TxGNN to test their robustness from perturbations. We found that TxGNN is robust with little performance drop against minor perturbations (from 10%-40%) of the signature vector, showing robustness. When under strong perturbations (50%-90%), the result is expected to be worse since it could no longer identify similar diseases. All evaluations are conducted with five random data splits (N=5). The mean performance is highlighted, while the 95% confidence intervals are represented by error bars.

### Supplementary Tables

| Method | KL | JS | Proximity | DSD | RGCN | HAN | HGT | BioBERT | TxGNN |
|---|---|---|---|---|---|---|---|---|---|
| Random | 0.50±0.01 | 0.49±0.01 | 0.53±0.04 | 0.49±0.02 | 0.84±0.02 | 0.87±0.18 | 0.72±0.20 | 0.81±0.02 | 0.91±0.02 |
| Zero-shot | 0.49±0.02 | 0.49±0.01 | 0.49±0.01 | 0.49±0.03 | 0.50±0.04 | 0.62±0.03 | 0.56±0.08 | 0.76±0.03 | 0.90±0.02 |
| Cell Proliferation | 0.51±0.01 | 0.50±0.01 | 0.47±0.00 | 0.51±0.00 | 0.47±0.00 | 0.65±0.04 | 0.61±0.10 | 0.83±0.0 | 0.92±0.00 |
| Mental Health | 0.50±0.02 | 0.50±0.02 | 0.44±0.00 | 0.50±0.01 | 0.55±0.01 | 0.61±0.03 | 0.56±0.04 | 0.64±0.0 | 0.89±0.00 |
| Cardiovascular | 0.49±0.01 | 0.50±0.02 | 0.50±0.00 | 0.50±0.02 | 0.41±0.00 | 0.63±0.05 | 0.60±0.05 | 0.66±0.0 | 0.64±0.00 |
| Anemia | 0.47±0.02 | 0.53±0.02 | 0.49±0.02 | 0.51±0.03 | 0.42±0.00 | 0.67±0.01 | 0.53±0.05 | 0.64±0.0 | 0.96±0.00 |
| Adrenal Gland | 0.48±0.05 | 0.50±0.02 | 0.48±0.02 | 0.50±0.03 | 0.38±0.00 | 0.59±0.04 | 0.54±0.10 | 0.64±0.0 | 0.99±0.00 |
| Autoimmune | 0.51±0.03 | 0.49±0.01 | 0.56±0.01 | 0.52±0.04 | 0.51±0.00 | 0.53±0.03 | 0.49±0.05 | 0.66±0.0 | 0.87±0.00 |
| Metabolic Disorder | 0.51±0.01 | 0.54±0.06 | 0.41±0.00 | 0.51±0.04 | 0.67±0.01 | 0.56±0.08 | 0.54±0.05 | 0.68±0.0 | 0.76±0.00 |
| Diabetes | 0.50±0.03 | 0.52±0.03 | 0.47±0.01 | 0.52±0.01 | 0.40±0.00 | 0.67±0.11 | 0.62±0.17 | 0.69±0.0 | 0.87±0.00 |
| Neurodegenerative | 0.50±0.03 | 0.51±0.03 | 0.49±0.01 | 0.51±0.02 | 0.84±0.00 | 0.71±0.10 | 0.60±0.06 | 0.57±0.0 | 0.95±0.00 |

**Table S1: Benchmarking models for predicting drug indications.** Reported is AUPRC.

| Method | KL | JS | Proximity | DSD | RGCN | HAN | HGT | BioBERT | TxGNN |
|---|---|---|---|---|---|---|---|---|---|
| Random | 0.49±0.01 | 0.50±0.01 | 0.56±0.00 | 0.49±0.01 | 0.77±0.02 | 0.84±0.00 | 0.63±0.19 | 0.58±0.02 | 0.82±0.01 |
| Zero-shot | 0.50±0.01 | 0.50±0.01 | 0.51±0.01 | 0.49±0.02 | 0.64±0.03 | 0.50±0.03 | 0.54±0.06 | 0.57±0.03 | 0.80±0.01 |
| Cell Proliferation | 0.49±0.01 | 0.49±0.01 | 0.50±0.00 | 0.50±0.00 | 0.54±0.00 | 0.51±0.04 | 0.47±0.03 | 0.78±0.0 | 0.87±0.00 |
| Mental Health | 0.50±0.01 | 0.50±0.00 | 0.50±0.00 | 0.50±0.01 | 0.67±0.00 | 0.44±0.02 | 0.56±0.04 | 0.54±0.0 | 0.79±0.00 |
| Cardiovascular | 0.49±0.00 | 0.49±0.00 | 0.51±0.00 | 0.50±0.00 | 0.63±0.00 | 0.47±0.02 | 0.51±0.02 | 0.56±0.0 | 0.72±0.00 |
| Anemia | 0.50±0.01 | 0.50±0.01 | 0.50±0.00 | 0.50±0.00 | 0.65±0.00 | 0.54±0.04 | 0.51±0.04 | 0.53±0.0 | 0.74±0.00 |
| Adrenal Gland | 0.49±0.01 | 0.50±0.01 | 0.48±0.00 | 0.49±0.01 | 0.75±0.00 | 0.46±0.02 | 0.55±0.08 | 0.47±0.0 | 0.88±0.00 |
| Autoimmune | 0.50±0.02 | 0.49±0.02 | 0.51±0.01 | 0.50±0.02 | 0.68±0.00 | 0.40±0.01 | 0.61±0.08 | 0.40±0.0 | 0.86±0.00 |
| Metabolic Disorder | 0.49±0.02 | 0.51±0.01 | 0.47±0.00 | 0.51±0.00 | 0.58±0.00 | 0.56±0.00 | 0.51±0.03 | 0.53±0.0 | 0.78±0.00 |
| Diabetes | 0.50±0.02 | 0.51±0.01 | 0.48±0.00 | 0.51±0.03 | 0.57±0.00 | 0.56±0.03 | 0.52±0.05 | 0.51±0.0 | 0.69±0.00 |
| Neurodegenerative | 0.51±0.02 | 0.51±0.02 | 0.56±0.02 | 0.51±0.02 | 0.71±0.00 | 0.42±0.01 | 0.55±0.04 | 0.60±0.0 | 0.78±0.00 |

**Table S2: Benchmarking models for predicting drug contraindications.** Reported is AUPRC.

| Disease area | Number of diseases | Number of indications | Number of contraindications |
|---|---|---|---|
| Diseases of cell proliferation | 183 | 854 | 1007 |
| Mental health diseases | 56 | 213 | 1038 |
| Cardiovascular diseases | 104 | 300 | 3131 |
| Diseases of anemia | 15 | 55 | 545 |
| Adrenal gland diseases | 6 | 33 | 303 |
| Autoimmune diseases | 18 | 75 | 319 |
| Metabolic disorders | 54 | 68 | 523 |
| Diabetes | 3 | 102 | 364 |
| Neurodegenerative diseases | 16 | 123 | 134 |

**Table S3: Statistics on disease-area-based dataset splits used to evaluate zero-shot drug repurposing.** Given all diseases in a given disease area, all indications and contraindications were removed from the dataset used to train machine learning models. Additionally, a fraction (5%) of the connections between biomedical entities to these diseases were removed from the therapeutics-centered knowledge graph. Disease-area splits were curated to evaluate model performance on diseases with limited molecular understanding and no existing treatments.

| | Accuracy | | Confidence | | Time | |
|---|---|---|---|---|---|---|
| User | Ours | Baseline | Ours | Baseline | Ours | Baseline |
| P1 | 1.00 | 0.50 | 4.25 | 2.50 | 69.00 | 16.50 |
| P2 | 0.75 | 1.00 | 2.75 | 1.25 | 32.50 | 13.00 |
| P3 | 0.75 | 0.75 | 3.00 | 3.25 | 49.25 | 18.50 |
| P4 | 0.25 | 0.25 | 3.25 | 2.00 | 68.25 | 17.50 |
| P5 | 0.50 | 0.50 | 4.00 | 3.25 | 39.50 | 14.00 |
| P6 | 1.00 | 0.75 | 3.50 | 2.00 | 57.75 | 16.75 |
| P7 | 0.75 | 0.50 | 2.50 | 2.25 | 86.25 | 25.75 |
| P8 | 1.00 | 0.50 | 4.75 | 4.25 | 72.00 | 26.50 |
| P9 | 0.75 | 0.50 | 4.25 | 2.50 | 45.90 | 21.30 |
| P10 | 1.00 | 0.50 | 3.25 | 1.25 | 85.20 | 18.70 |
| P11 | 1.00 | 0.25 | 3.00 | 2.00 | 58.50 | 17.50 |
| P12 | 0.75 | 0.50 | 4.00 | 2.00 | 35.60 | 14.30 |
| Mean | 0.79 | 0.54 | 3.54 | 2.38 | 58.31 | 18.36 |
| STD | 0.23 | 0.21 | 0.70 | 0.86 | 18.28 | 4.28 |

**Table S4: Accuracy, confidence, and time reported for participants in the human evaluation study of TxGNN.**

| | Accuracy | | Confidence | | Time | |
|---|---|---|---|---|---|---|
| Task | Ours | Baseline | Ours | Baseline | Ours | Baseline |
| T1 | 1.00 | 1.00 | 4.67 | 3.33 | 53.00 | 23.40 |
| T2 | 0.33 | 0.67 | 3.00 | 3.67 | 54.00 | 26.33 |
| T3 | 0.67 | 1.00 | 2.67 | 2.00 | 61.33 | 11.60 |
| T4 | 1.00 | 0.33 | 3.00 | 2.33 | 60.67 | 21.33 |
| T5 | 1.00 | 0.00 | 4.67 | 2.67 | 48.33 | 16.67 |
| T6 | 1.00 | 0.00 | 4.67 | 3.00 | 56.67 | 15.00 |
| T7 | 0.67 | 0.33 | 3.67 | 1.33 | 57.00 | 12.67 |
| T8 | 1.00 | 0.33 | 3.67 | 2.33 | 67.00 | 12.33 |
| T9 | 0.67 | 0.33 | 3.00 | 3.00 | 51.33 | 19.00 |
| T10 | 0.67 | 0.33 | 5.00 | 1.67 | 57.33 | 23.67 |
| T11 | 1.00 | 0.67 | 1.67 | 1.33 | 80.67 | 21.00 |
| T12 | 1.00 | 0.67 | 1.67 | 2.00 | 75.33 | 15.67 |
| T13 | 1.00 | 0.33 | 1.67 | 2.00 | 75.33 | 18.67 |
| T14 | 0.67 | 1.00 | 3.33 | 1.67 | 66.47 | 17.33 |
| T15 | 1.00 | 1.00 | 3.33 | 1.33 | 59.67 | 19.33 |
| T16 | 0.67 | 0.67 | 3.67 | 4.33 | 51.27 | 19.73 |
| Mean | 0.83 | 0.54 | 3.33 | 2.37 | 60.96 | 18.36 |
| STD | 0.21 | 0.34 | 1.08 | 0.89 | 9.58 | 4.27 |

**Table S5: Accuracy, confidence, and time for every task in the human evaluation study of TxGNN.**

| Racial group | Number of patients | Percent (%) |
|---|---|---|
| Asian | 60,041 | 4.7 |
| Black | 162,102 | 12.7 |
| White | 534,305 | 42.0 |
| Unknown | 241,998 | 19.0 |
| Other | 273,639 | 21.5 |
| Total number of patients | 1,272,085 | 100.0 |

**Table S6: Demographics of the electronic health record (EHR) dataset at Mount Sinai Health System in New York City.** The EHR dataset was used to perform an independent evaluation of TxGNN's novel drug repurposing predictions.

| Drug name | Active ingredient | Disease | Approval | FDA number | Orphan | TxGNN | Percentile |
|---|---|---|---|---|---|---|---|
| Vabysmo | Faricimab | Macular degeneration | 01/28/2022 | BLA761235 | No | 0.938 | 2.25% |
| Welireg | Belzutifan | von Hippel-Lindau disease | 08/13/2021 | NDA215383 | Yes | 0.720 | 4.11% |
| Mounjaro | Tirzepatide | Type 2 diabetes mellitus | 05/13/2022 | NDA215866 | No | 0.286 | 12.50% |
| Ztalmy | Ganaxolone | CDKL5 disorder | 03/18/2022 | NDA215904 | Yes | 0.335 | 18.73% |
| Leqvio | Inclisiran sodium | Familial hypercholesterolemia | 12/22/2021 | NDA214012 | No | 0.301 | 19.32% |
| Tezspire | Tezepelumab-ekko | Asthma | 12/17/2021 | BLA761224 | No | 0.233 | 32.41% |
| Vtama | Tapinarof | Psoriasis | 05/23/2022 | NDA215272 | No | 0.261 | 32.70% |
| Adbry | Tralokinumab | Atopic dermatitis | 12/27/2021 | BLA761180 | No | 0.040 | 50.37% |
| Vonjo | Pacritinib citrate | Myelofibrosis | 02/28/2022 | NDA208712 | Yes | 0.011 | 63.14% |
| Livtencity | Maribavir | Cytomegalovirus infection | 11/23/2021 | NDA215596 | Yes | 0.033 | 66.37% |

**Table S7: Evaluation of TxGNN on recent FDA-approvals.** To demonstrate that TxGNN was not driven by confirmation bias from indications and contraindications already present in the knowledge graph, we considered ten therapies that were approved by the FDA after TxGNN's dataset and model development were completed (June 2021). None of these therapies had direct relationships between their drug-disease nodes in the TxGNN dataset. We then asked TxGNN to make predictions for those diseases without any prompting for the recently approved drug and observed that TxGNN consistently ranked newly introduced drugs highly, with the recently approved drugs found in the first third (30.19%) of the full-length prediction list on average. Occasionally, TxGNN ranked the approved drug in the top 5% of therapeutic candidates, such as faricimab to treat macular degeneration (top 2.25%) and belzutifan to treat von Hippel-Lindau disease (top 4.11%). In one case, TxGNN ranked maribavir in the bottom two-thirds of the prediction list for treating cytomegalovirus infections in patients post-transplantation, likely because TxGNN's knowledge graph did not contain information about the host-pathogen interactions that inform the treatment pathway.

| Node Type | Count | Percent (%) |
|---|---|---|
| Biological process | 28,642 | 22.1 |
| Protein | 27,671 | 21.4 |
| Disease | 17,080 | 13.2 |
| Phenotype | 15,311 | 11.8 |
| Anatomy | 14,035 | 10.8 |
| Molecular function | 11,169 | 8.6 |
| Drug | 7,957 | 6.2 |
| Cellular component | 4,176 | 3.2 |
| Pathway | 2,516 | 1.9 |
| Exposure | 818 | 0.6 |
| Total number of nodes | 129,375 | 100.0 |

**Table S8: Statistics on nodes in the medical knowledge graph (KG).**

| Relation | Count | Percent (%) |
|---|---|---|
| Anatomy – Protein (present) | 3,036,406 | 37.5 |
| Drug – Drug | 2,672,628 | 33.0 |
| Protein – Protein | 642,150 | 7.9 |
| Disease – Phenotype (positive) | 300,634 | 3.7 |
| Biological process – Protein | 289,610 | 3.6 |
| Cellular component – Protein | 166,804 | 2.1 |
| Disease – Protein | 160,822 | 2.0 |
| Molecular function – Protein | 139,060 | 1.7 |
| Drug – Phenotype | 129,568 | 1.6 |
| Biological process – Biological process | 105,772 | 1.3 |
| Pathway – Protein | 85,292 | 1.1 |
| Disease – Disease | 64,388 | 0.8 |
| Drug – Disease (contraindication) | 61,350 | 0.8 |
| Drug – Protein | 51,306 | 0.6 |
| Anatomy – Protein (absent) | 39,774 | 0.5 |
| Phenotype – Phenotype | 37,472 | 0.5 |
| Anatomy – Anatomy | 28,064 | 0.3 |
| Molecular function – Molecular function | 27,148 | 0.3 |
| Drug – Disease (indication) | 18,776 | 0.2 |
| Cellular component – Cellular component | 9,690 | 0.1 |
| Phenotype – Protein | 6,660 | 0.1 |
| Drug – Disease (off-label use) | 5,136 | 0.1 |
| Pathway – Pathway | 5,070 | 0.1 |
| Exposure – Disease | 4,608 | 0.1 |
| Exposure – Exposure | 4,140 | 0.1 |
| Exposure – Biological process | 3,250 | $<$0.1 |
| Exposure – Protein | 2,424 | $<$0.1 |
| Disease – Phenotype (negative) | 2,386 | $<$0.1 |
| Exposure – Molecular function | 90 | $<$0.1 |
| Exposure – Cellular component | 20 | $<$0.1 |
| Total number of edges | 8,100,498 | 100.0 |

**Table S9: Statistics on edges in the medical knowledge graph (KG).**
